# Lesions causing room tilt illusion link a cerebello-cingulate brain network to verticality perception

**DOI:** 10.64898/2026.10.26.26365825

**Authors:** Lorenzo Pini, Karoline Steinmetz, Charlie Weige Zhao, Arun Garimella, Zoé Dary, Frederic L.W.V.J. Schaper, Fabrice Bartolomei, Stanislas Lagarde, Jean Laurens, Nico Sollmann, David S. Zee, Andreas Zwergal, Marianne Dieterich, Bernhard Baier, Christophe Lopez, Michael D. Fox, Julian Conrad, Maximilian U. Friedrich

**Affiliations:** Department of Neurology, University Hospital Ulm, Ulm, Germany; Department of Diagnostic and Interventional Radiology, University Hospital Ulm, Ulm, Germany; Department of Nuclear Medicine, University Hospital Ulm, Ulm, Germany; LMU Department of Neurology & German Center for Vertigo and Balance Disorders (DSGZ), LMU University Hospital, LMU Medizin, Munich, Germany; Center for Brain Circuit Therapeutics, Brigham & Women’s Hospital, Boston, MA, USA; Harvard Medical School, Boston, MA, USA; Department of Neurosurgery, Mass General Brigham, Harvard Medical School, Boston, MA, USA; Epileptology and Cerebral Rhythmology, APHM, Timone Hospital, Marseille, France; Aix Marseille University, Inserm, Institute de Neuroscience des Systèmes, Marseille, France; Ernst Strüngmann Institute for Neuroscience, Frankfurt, Germany; Department of Diagnostic and Interventional Neuroradiology, School of Medicine and Health, Technical University of Munich, TUM Klinikum Rechts der Isar, Munich, Germany; TUM-Neuroimaging Center, TUM Klinikum Rechts der Isar, Technical University of Munich, Munich, Germany; Department of Neurology, Johns Hopkins Hospital, Baltimore, MD, USA; Spinal Cord Injury Center, BG-Klinik Ludwigshafen, Ludwigshafen, Germany; Center for Research in Psychology and Neuroscience (CRPN), Aix Marseille University and Centre National de la Recherche Scientifique (CNRS), Marseille, France; Division of Neurodegeneration, Department of Neurology, Medical Faculty Mannheim, Heidelberg University, Mannheim, Germany

**Keywords:** Verticality perception, Vestibular system, Multisensory processing, Network mapping, Neuroimaging

## Abstract

Verticality perception is the sense of one’s body orientation relative to gravity. It can be disrupted by neurological diseases or during aviation and space flight, resulting in falls and accidents. This perception is thought to arise from integration of vestibular and visual signals, yet where this multisensory computation is spatially implemented in the human brain remains unresolved. Room tilt illusion (RTI), in which the visual environment abruptly reorients relative to gravity (typically by 90° or 180°), has been proposed as a model disorder for studying verticality perception. Here, we leverage focal brain lesions (predominantly ischemic or hemorrhagic strokes) causing RTI to localize this multisensory computation. We assembled a total of 37 patients with brain lesions causing RTI from a systematic literature search and local cases across two centers. Clinical data were tabulated and lesions were analyzed using multimodal voxel- and connectome-based lesion mapping methods. Lesion findings were further tested against an independent dataset of 509 direct electrical stimulation sites associated with vestibular or non-vestibular percepts. The RTI lesions were distributed across cortical and subcortical locations, with the cerebellum representing the most frequent lesion site (49%). Despite anatomical heterogeneity, lesion connectivity converged on two hub regions: one in the posterior cerebellum and another in the mid-cingulate cortex. This pattern was robust to methodological variations, separation of RTI subtypes, and exclusion of cerebellar lesions. This pattern was also specific to RTI when compared against 185 phenotypically matched lesions from patients with milder disturbances of verticality perception and 1,110 lesions associated with 27 other neuropsychiatric syndromes. Finally, independent direct electrical stimulation data converged on both lesion-derived RTI hubs (mid-cingulate cortex: g=0.62, p<.001; posterior cerebellum: g=0.34, p=.014, 10,000 label permutations). In conclusion, anatomically diverse RTI lesions localize to a specific cerebello-cingulate network, providing an anatomical framework for verticality perception in the human brain. Our findings generate testable hypotheses for experimentally modulating or protecting verticality perception in neurological patients or pilots.

## INTRODUCTION

Gravity shapes the physical world and how organisms interact with it. Across species, perceiving its direction is a fundamental neural computation that underpins gaze, posture, and spatial orientation. In humans, experimental observations indicate that spatial orientation relative to gravity depends on the reliability-weighted integration of vestibular, visual, and other sensory cues into a coherent, global estimate of verticality^1–3^. Errors in the underlying computations have real-world consequences, contributing to falls on the ground^4^ and potentially fatal aviation mishaps due to spatial disorientation in pilots^5^.

Disordered verticality perception is also a frequent and disabling feature of neurological conditions such as stroke, Parkinsonism, or vestibular disorders, where it contributes to poor clinical outcomes, mostly related to falls and their sequelae^4,6,7^. Notably, verticality perception can be modulated through both invasive and non-invasive neuromodulation, suggesting that it may represent a tractable network-level target, both in neurological contexts^8–12^ and for mitigating spatial disorientation in aviation^13^. Although the multisensory computations underlying verticality perception have been examined in detail through psychophysical experiments^1,3,14^, where these computations are implemented and spatially encoded in the human brain remains incompletely resolved^15^.

Room tilt illusion (RTI) is considered a unique model disorder to localize the neural substrates mediating verticality perception, as proposed by Alberts and colleagues^14^. During RTI, patients experience an abrupt and extreme rotation of the entire visual environment relative to gravity, most commonly by 90° or 180°, while the scene itself remains internally coherent and most other aspects of perception intact^16,17^. In contrast to the more common, milder disturbances of verticality perception (e.g., tilts of the subjective visual vertical, SVV^18–20^) that produce a continuously tilted percept, RTI appears quantized, collapsing onto a small set of stable spatial configurations^16^. Albeit also described in peripheral vestibular disorders, RTI is most commonly caused by brain lesions^16,21^. These lesions have been reported across remarkably distributed locations, spanning cortical and subcortical regions linked to vestibular, visual, and multisensory processing^16,22^. This observation underpins the long-standing hypothesis that RTI reflects a brain network disorder characterized by a transient mismatch between vestibular and visual coordinate maps of space^23,24^. Although the clinical and imaging features of RTI have been systematically reviewed^16^, RTI-causing lesions have not been systematically analyzed at the voxel- and network level, leaving unresolved whether shared network-level substrates exist, and if so, where they are localized. Lesion network mapping provides a means of identifying shared circuitry across anatomically heterogeneous lesions, and it has already revealed convergent networks underlying specific disorders of higher-order vestibular, visual, and multisensory processing^25–27^.

Based on a combined local and literature-based cohort of 37 lesion-induced RTI cases, we applied functional and structural connectivity analyses across independent lesion and brain stimulation datasets, each modeling complementary aspects of verticality and vestibular perception. We show that anatomically heterogeneous RTI lesions converge onto a specific cerebello-cingulate network that is embedded within the canonical central vestibular network, but is dissociable from the network underlying milder disturbances of verticality perception. Together, these findings can provide a causal neuroanatomical framework for localizing multisensory computations in the human brain, generating testable hypotheses for understanding and modulating human verticality perception.

## METHODS

The whole methodological workflow is depicted in Figure 1. See supplementary information for additional details. This retrospective study was approved by the institutional review board of Brigham and Women’s Hospital, Boston, MA, USA (#2020P002987), and was deemed exempt from review at the institutional review board of the University Hospital Ulm, Germany.

**Figure 1.**
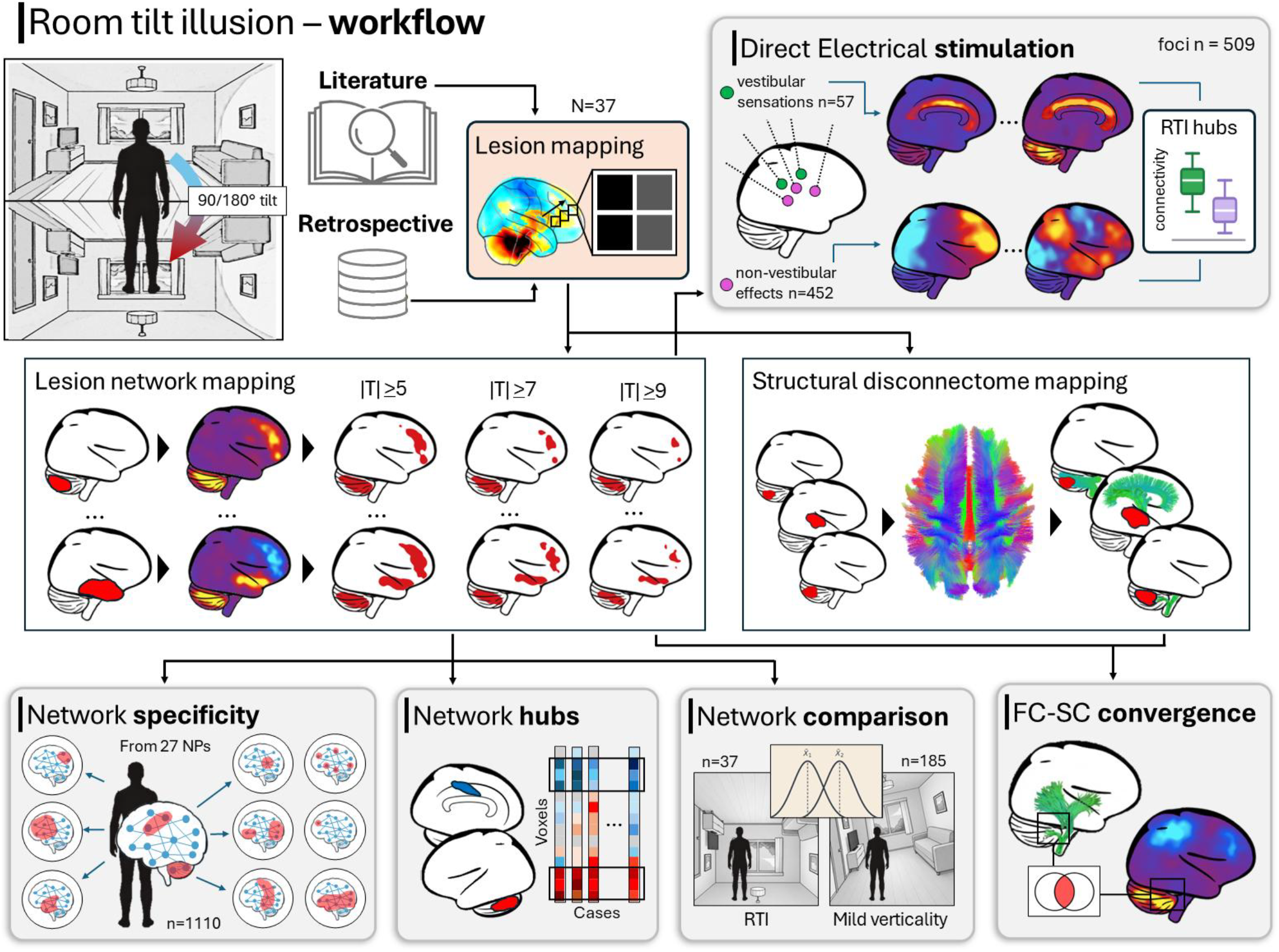
Methodological workflow. Room tilt illusion (RTI) cases were collected from the literature and retrospective local cases. Each lesion was mapped onto a normative functional and structural connectome to obtain its connectivity maps. The following analyses were performed: i) functional connectivity maps were compared against lesion networks of n=27 other lesion-induced neuropsychiatric disorders; ii) network maps were combined across cases to find network hubs; iii) RTI networks were compared with cases with mild disturbances of verticality; iv) the RTI network was compared with direct electrical stimulation sites. Abbreviations: RTI: Room tilt illusion; FC: functional connectivity; SC: structural connectivity. NPs: neuropsychiatric disorders.

### Syndromic definition and case collection

Disturbances of verticality perception span a clinical spectrum defined by the magnitude of the perceived deviation from true vertical. Common manifestations, such as the ocular tilt reaction (OTR) or isolated pathological deviations of the SVV, are typically associated with tilt angles above 2.5° but well below 30°^28,29^. In contrast, RTI is an extreme misperception of verticality that is characterized by tilts of the visual environment reaching 45°, 60°, 90°, or 180°, most commonly in the roll plane. For the purpose of this study and in line with prior literature, we operationally defined the latter category as RTI, irrespective of the heterogeneous terminology used in the original reports (e.g., “upside-down vision”, “environmental tilt illusion”, or “reversal of vision metamorphopsia”)^16^.

The study was based on two independent lesion datasets: retrospective local cases and cases identified through a systematic literature search following the Preferred Reporting Items for Systematic Reviews and Meta-Analyses (PRISMA) guidelines^30^. Searches were performed on PubMed in August 2024 with the following search terms: (1) OTR or SVV deviation: (“skew deviation” OR “ocular tilt reaction” OR “visual vertical”) AND (“case report” OR “case series” OR “case”); (2) RTI: ((“visual inversion” OR “visual rotation” OR “visual tilt” OR ((“vision” OR “visual”) AND (“180 degrees” OR “upside down” OR “upside-down”))) AND (“case” OR “cases”) AND (“CT” or “MRI” or “imaging” or “neuroimaging”)) OR “room tilt illusion” OR “environmental tilt illusion” OR (“reversal” AND “vision” AND “metamorphopsia”); (3) OTR, SVV deviation, or RTI cases: “vestibular cortex” AND (“case” OR “case series” OR “case report”).

For RTI, the literature search identified 33 cases from 27 studies^16,21,31^. An additional four local and unpublished cases across two institutions were included (Supplementary Figure 1). Cases were included if they met the following criteria: (1) presence of an extreme (>30°) misperception of verticality, operationally defined as RTI; (2) presence of a focal brain lesion temporally associated with symptom expression; and (3) availability of clinical neuroimaging of sufficient quality for accurate lesion segmentation.

For the secondary, comparative analyses, a dataset of 185 lesions causing milder disturbances of verticality perception (≤30°) was assembled. This comprised 89 cases of lesion-induced ocular tilt reaction and/or documented SVV deviations, identified through an independent PRISMA-based literature search (Supplementary Figure 2). In addition, a local multi-institutional convenience sample comprising 96 lesion cases similarly defined by SVV deviations (≥2.5°, as assessed with paradigms such as a visual dome or the “bucket test”)^32^, was collected. These cases have been collected at the LMU Munich and University of Mainz, as reported previously^19,20,33^.

### Lesion network mapping

To approximate the brain-wide network connections of a given lesion location, we used the most recent and validated iteration of the lesion network mapping methodology^26,34–36^, which encompasses the following steps.

#### Lesion segmentation

Lesion locations were manually transferred from published figures or local neuroimaging documentation onto a reference brain atlas in standard space (Montreal Neurological Institute 152 [MNI152] with 2 mm isotropic resolution) using MRIcroGL (www.nitrc.org/projects/mricrogl), in line with previous studies^26,34,37^. To ensure reproducibility, two assessors (MUF, CWZ) segmented all independently for lesions subsequent comparisons, consistent with current best practice recommendations^38^. Inter-assessor agreement was quantified as the voxel-wise Pearson correlation between the lesion network maps derived from the two assessors’ independent segmentations. For cases derived from the literature, lesion locations were traced on all published neuroimaging slices, irrespective of number or orientation, and each lesion was traced to its full visible extent on every available slice. Of note, previous work demonstrated that two-dimensional (2D) lesion traces are sufficient for lesion network mapping^25,39^. For the local cases, the full clinical imaging dataset was available, and lesions were manually segmented in their entire three-dimensional extent.

#### Lesion mapping

To integrate the anatomical distribution of lesion locations associated with different severities of verticality misperception, lesion overlap maps were generated for lesions causing RTI and lesions associated with milder SVV tilts (≤30°). Lesion masks were binarized and combined to create voxel-wise overlap maps representing the fraction of cases involving each voxel. To identify neuroanatomical regions preferentially associated with RTI compared with milder SVV tilts, we performed voxel-wise lesion-symptom mapping (VLSM) using the MATLAB R2018a package NiiStat, as previously described^37,40^. Voxel-wise Liebermeister tests were used to contrast lesions associated with RTI against those associated with milder SVV tilts (≤30°). Lesion volume was included as a nuisance covariate to account for potential effects of lesion size, as well as a minimum overlap of 5, 10, and 20 lesions, in line with our previous study^37^. Statistical significance was assessed using family-wise error (FWE) correction based on 2,000 Freedman–Lane permutations, as implemented in NiiStat.

#### Using the human functional connectome to compute lesion networks

Functional connectivity between each lesion and the rest of the brain was estimated using a normative resting-state functional connectome derived from 1,000 healthy adults (50% female; age range 18-35 years) from the Brain Genomics Superstruct Project (GSP), acquired at Harvard University and Massachusetts General Hospital using a 3-T Siemens Magnetom TrioTim magnetic resonance imaging (MRI) scanner with a 12-channel phased-array head coil^41,42^. Structural images consisted of high-resolution T1-weighted scans (1.2 mm isotropic resolution; repetition time [TR] = 2200 ms; inversion time [TI] = 1100 ms; flip angle = 7°; acquisition matrix = 192 × 192 with parallel imaging (GRAPPA) acceleration factor = 4), whereas resting-state functional images were acquired using gradient-echo echo-planar imaging (47 interleaved slices (foot-head); slice thickness = 3 mm with no interslice gap; TR = 3000 ms; TE = 30 ms; flip angle = 85°; 120 usable volumes per run following removal of the first four volumes). Participants completed one or two resting-state runs (72.6% completed two runs). Standard preprocessing included regression of motion, cerebrospinal fluid, white matter and global signal fluctuations, as described previously^41,42^.

The resulting dataset provided a normative estimate of whole-brain functional connectivity, allowing the connectivity profile of each lesion location to be inferred in a common reference space. For each lesion mask, the mean resting-state time course was correlated with every other brain voxel across all subjects, yielding a voxel-wise connectivity map that was converted into a T-statistic, thus reflecting the consistency of connectivity across the normative cohort. Positive and negative values indicated regions showing correlated and anticorrelated spontaneous blood-oxygenation-level-dependent (BOLD) fluctuations with the lesion location, respectively^43^. To evaluate the robustness of our findings to connectome preprocessing choices, the complete connectivity analysis was repeated using both currently established preprocessing pipelines for the GSP connectome^41,42,44^.

### Lesion network overlap

In line with validated pipelines for lesion network mapping, we thresholded the lesion network maps at |T|≥ 5 (corresponding to a voxel-wise FWE-corrected p< .05) to assess the consistency of lesion connection in the RTI cohort, creating binarized maps of brain regions significantly connected to the lesion location. We repeated this process at |T|≥ 7 (voxel-wise FWE-corrected p< 10^-6^), and |T|≥ 9 (voxel-wise FWE-corrected p< 10^-11^) to verify robustness against threshold variations. Finally, we overlaid the binarized lesion network maps to identify peak connectivity regions across the anatomically heterogeneous lesions causing RTI. As an alternative, threshold-free measure of network consistency^45^, voxel-wise connectivity values from all individual lesion network maps were entered into a one-sample t-test against a null hypothesis of zero mean connectivity. This analysis retains continuous voxel-wise connectivity values, thereby providing a complementary measure of convergence across lesions.

### Specificity testing

As lesion network overlap alone is insufficient to detect connections specific to a group of lesions of interest^35^, we conducted two independent specificity analyses. First, we contrasted the unthresholded lesion network maps from the 37 RTI lesions with 1,110 lesion network maps derived from lesions causing 27 other neuropsychiatric disorders as previously reported^26^, including other central visual and/or perceptual disorders (e.g., Alice in Wonderland syndrome^26^, hallucinations^46^, prosopagnosia^47^, or cortical blindness^25,48^). Second, to test for connections specific to different degrees of misperceptions of verticality, we performed a similar two-sample t-test comparing RTI to lesion network maps associated with milder disturbances of verticality perception as a control cohort phenotypically matched for disturbances in verticality perception (n = 185).

All of these tests accounted for lesion size as a nuisance variable and were implemented using open software for statistical analysis of neuroimaging data (FSL’s permutation analysis of linear models, PALM^49^). Results were corrected for multiple comparisons using voxel-based FWE correction in FSL-PALM. To characterize the magnitude of group differences within the data-driven RTI network hubs, we additionally extracted mean connectivity values from each individual lesion network map and calculated Hedges’ g. Group differences were also assessed using non-parametric label permutation testing with 10,000 permutations^35,50^. Because the regions of interest (ROIs) were derived from the RTI dataset itself, these ROI-based analyses were considered descriptive and complementary to the whole-brain voxel-wise inference rather than an independent test of specificity.

An analogous ROI-based permutation framework was applied to connectivity maps derived from direct electrical stimulation (DES) sites. This was done to test whether the lesion-derived RTI network hubs were preferentially connected to stimulation sites eliciting vestibular compared with non-vestibular percepts. As the stimulation and lesion datasets in that case were independent, this analysis provided an independent test of the functional specificity of the RTI hubs. Label permutation testing was used to assess whether these group differences exceeded those expected by chance, in line with current methodological recommendations emphasizing permutation-based specificity testing in connectome-based brain mapping studies^35,50^.

### Deriving a lesion-based network for RTI

A conjunction map was created by combining the lesion network overlap map and the specificity map using the logical “and” operation in Python 3.0’s nibabel library. This combination identified brain regions consistently (i.e., connected in ≥89% of cases) and specifically (i.e., regions with FWE-corrected p< .05 in the specificity analysis) connected to RTI lesions. We will refer to the areas resulting from this conjunction analysis as RTI “network hubs”. As an exploratory robustness analysis, we additionally used permutation-based null distributions to estimate false-positive rates across a range of lesion network overlap, specificity, and conjunction thresholds.

We compared the location of these hubs to advanced structural brain atlases including the brainnetome atlas^51^, the Spatially Unbiased Infra-tentorial Template (SUIT) cerebellum atlas^52^, and the Harvard-Oxford cortical atlas^53^. In the case of cortical regions of interest, hub locations and atlas boundaries were compared by overlaying both on a high-resolution MRI template (400 μm isotropic resolution, Juelich EBrains) in FSLeyes^54^, while in the case of cerebellar regions of interest, neuroimaging findings were projected onto SUIT’s cerebellar flatmap visualization^55^, as per the instructions supplied with the SUITPy package in Python 3.0 (https://suitpy.readthedocs.io/en/latest/flatmap.html). By definition, positive connectivity to the positive RTI hub and negative connectivity to the negative RTI hub defined a distributed brain network encompassing the heterogeneous lesion locations associated with RTI. To visualize this network, we computed functional connectivity with the two RTI network hubs, thresholded each connectivity map at |T|= 5, and identified all voxels that satisfied the above criteria.

### Direct electrical stimulation network mapping

To further test whether the lesion-derived RTI network hubs participated in vestibular processing, we utilized a convergent brain mapping approach by analyzing an independent dataset of DES sites associated with vestibular and non-vestibular percepts, as previously reported^25,56^. Cases were identified from a retrospective analysis of 354 patients with medication-resistant focal epilepsy, who underwent presurgical evaluation with stereo-electroencephalography (SEEG) and electrical brain stimulation^57,58^.

Specifically, DES was performed between two adjacent contacts of the SEEG electrodes (inter-contact space = 0.5 mm) at 50 Hz (pulse width: 0.5–1 ms; train duration: 3–8 s) and at 1 Hz (pulse width: 1–2 ms; train duration: 20–60 s). The intensity was increased progressively by 0.2 mA steps from 0.2 mA (maximum: 5 mA, rarely exceeding 3 mA). Patients were blinded to stimulation timing and systematically asked for subjective experiences^57^.

The DES sites were modelled as 4 mm spheres centered at the midpoint between the two stimulated contacts. Brain-wide connectivity was analyzed using the same normative functional connectome and processing pipelines. Vestibular sites comprised DES locations eliciting sensations of vertigo, dizziness, rotations, tilts, and swaying (n = 57). All remaining DES sites eliciting motor, autonomic, language, affective, perceptual (e.g., somatosensory, visual, or auditory) effects as well as sites producing no effect served as controls (n = 452). To identify connections preferentially associated with vestibular stimulation sites, voxel-wise two-sample t-tests were performed using FSL-PALM, comparing vestibular and non-vestibular stimulation-derived connectivity maps. As an independent validation analysis, mean connectivity within lesion-derived RTI network hubs was compared between vestibular and non-vestibular DES sites using non-parametric label permutation testing (10,000 permutations).

### Disconnectome mapping

We generated structural disconnectome maps for each lesion using BCBtoolkit^59^. To this end, a normative structural connectome from 7-T diffusion MRI data of n = 178 healthy controls was used, as previously described^37,59^. This connectome reflected the inter-individual variability of fiber pathways in the healthy human brain. In line with previous work, the individual tractography maps associated with each lesion were then binarized at >0.5 (reflecting disconnection in the majority of connectome subjects) and collapsed across individuals, resulting in a disconnectome map for each lesion^37,59^. These maps reflected the probability of anatomical disconnection between the lesion location and each other brain voxel in the normative fiber connectivity dataset. To test for disconnections specific to either subgroup of verticality misperceptions (i.e., lesions causing RTI and those causing ≤30° verticality deviations), a two-sample t-test, accounting for lesion size as a nuisance variable, was implemented in FSL-PALM^49^. All results were corrected for multiple comparisons using voxel-based FWE correction with a significance level set at p< .05.

### Statistical computations and visualization

Spatial correlations, permutation testing, and combination of lesion network maps (e.g., for conjunction map) were conducted using Python (version 3; https://www.python.org) with Pandas, NumPy, Nibabel, and SciPy packages. Data distributions (e.g., demographic factors, clinical case features) were determined using Shapiro-Wilk testing or inspection of interquantile (“Q-Q”) plots. Depending on the data distribution, parametric (t-test) or non-parametric (Mann–Whitney U) tests were used for continuous variables, and Fisher’s exact test for categorical variables. Results from voxel-wise analyses were corrected for multiple comparisons using voxel-wise FWE correction in FSL-PALM. Data are reported as mean ± standard deviation or median and interquartile range (IQR). The significance level was set at p< .05. Main findings are visualized on the Ebrains BigBrain template normalized to MNI152 space^60^, or on a cerebellar flatmap visualization as implemented in Python using the SUITPy package^55^.

## RESULTS

### Demographic and clinical characteristics of RTI cases

Combining local cases and our systematic literature search yielded a total of 37 cases of RTI attributable to focal brain lesions (14 right, 15 left, 8 bilateral, Supplementary Table 1, Supplementary Figure 3). Of these, 22 (59%) presented with a 180° tilt/inversion, 14 (38%) with a 90° tilt, and one case (3%) with a 45° tilt. Across 180° and 90° subgroups, vascular etiologies predominated, accounting for 64% of 180° and 93% of 90° cases, followed by inflammatory (14% vs. 0%) and other causes (22% vs. 7%). Patients with 180° RTI were significantly younger than those with 90° RTI (p = 0.035). Conversely, vestibular symptoms and signs, including vertigo or dizziness (74% vs. 64%), the ocular tilt reaction (29% vs. 57%), and nystagmus (69% vs. 50%), were common in both RTI subgroups, as was ataxia (65% vs. 77%). Among cases with documented nystagmus, gaze-evoked nystagmus occurred at similar frequencies in 180° and 90° RTI (25% vs. 40%), whereas spontaneous nystagmus was reported exclusively in three 180° RTI cases. Nevertheless, nystagmus subtype did not differ significantly between groups (all Fisher’s exact p> 0.2). All RTI episodes were generally transient, with a median duration of 5 minutes (IQR 29.0; range 0.1 – 4320 minutes; duration not reported in 8/37 cases). Among cases with available contextual information, episodes occurred almost exclusively indoors (20/21 cases, 95%). Detailed cohort characteristics are summarized in Table 1.

**Table 1.** Demographic and clinical characteristics of patients grouped by degree of RTI. Patients affected by 180° RTI were significantly younger, and causal lesions were more frequently subcortical. Conversely, there were no significant differences in sex, lesion etiology and associated clinical features, which mainly represented signs of central vestibular dysfunction, and ataxia.

| Characteristic | 180° RTI (n= 22) | 90° RTI (n= 14) | p-value* |
| --- | --- | --- | --- |
| **Demographics** |  |  |  |
| Age, years, median (IQR) | 48 (33–60) | 63 (57–66) | **0.035** |
| Male sex, n (%) | 15 (68) | 11 (79) | ns |
| **Etiology, n (%)** |  |  |  |
| Vascular (ischemic/hemorrhagic) | 14 (64) | 13 (93) | ns |
| Inflammatory | 3 (14) | 0 (0) | ns |
| Other | 5 (22) | 1 (7) | ns |
| **Lesion topography, n (%)** |  |  |  |
| Subcortical | 13 (59) | 14 (100) | **0.006** |
| Cortical | 9 (41) | 0 (0) |  |
| **Clinical features, n/N reported (%)** |  |  |  |
| Vertigo or dizziness | 14/19 (74) | 9/14 (64) | ns |
| Nystagmus | 11/16 (69) | 5/10 (50) | ns |
| Ocular tilt reaction | 2/7 (29) | 4/7 (57) | ns |
| Ataxia | 11/17 (65) | 10/13 (77) | ns |

\* Comparison between 180° and 90° subgroups; Mann-Whitney U-test for age, Fisher's exact test for categorical variables. IQR, interquartile range. Clinical feature percentages are calculated with denominators restricted to cases with reported status.

### Lesion topography

Across the RTI cohort, lesions were distributed throughout various brain regions, most frequently involving the cerebellum (18 cases, 49%), pons (5 cases, 14%), occipital cortex (5 cases, 14%), midbrain (3 cases, 8%), and medulla (3 cases, 8%). Thalamic or temporal cortex involvement each was observed in 2 cases (5%), while the frontal cortex was affected in a single case (3%). Within the cerebellum, posterior lobules (lobules VIII–X, Crus II) were involved in the majority of cases (13/18, 72%), while anterior/intermediate regions (lobules IV– VI) were less frequently affected (6/18, 33%), with overlapping involvement in one case.

Lesions causing RTI were commonly located in subcortical structures (73%, 27/37) and equally distributed across both brain hemispheres (15 left, 14 right, 8 bilateral, Supplementary Figure 3). Cortical lesions were significantly more common in 180° RTI (9/ 22 vs. 0/ 14, p = .006). Consistent with this regional heterogeneity, descriptive voxel-wise overlap was low, with a maximum of 4/ 37 lesions (11%) located in the left paramedian aspect of cerebellar lobule IX (Supplementary Figure 4).

### Lesions causing different degrees of RTI converge on one common brain network

Despite anatomical heterogeneity (Figure 2), lesion locations causing RTI were consistently functionally connected to two principal hub regions, one in the posterior cerebellum and another in the cingulate cortex (Figure 3A-C).

**Figure 2.**
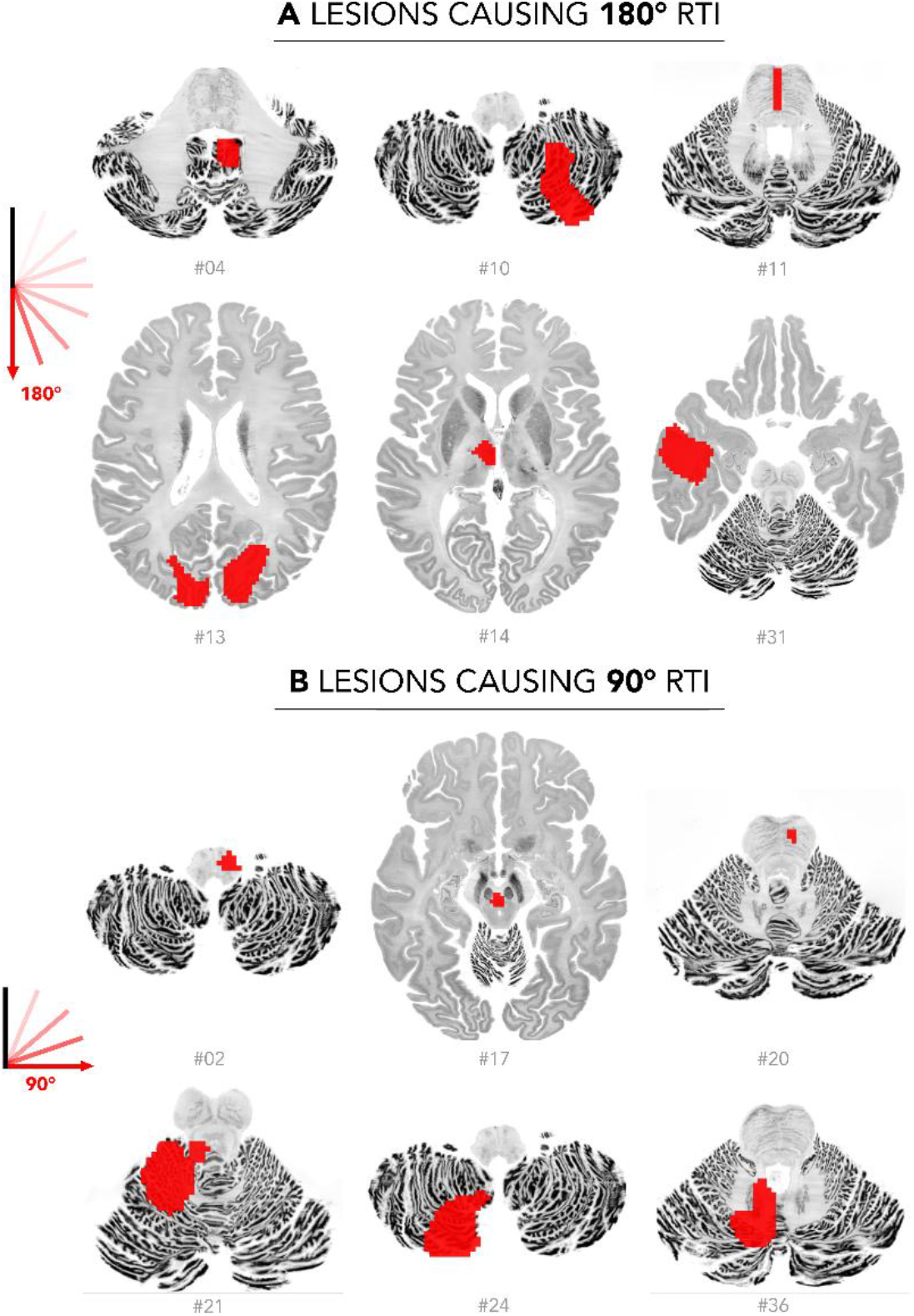
Lesions causing RTI are neuroanatomically heterogeneous. A) Lesions causing 180° tilts / inversions and B), lesions causing 90° tilts, all depicted in radiological convention.

**Figure 3.**
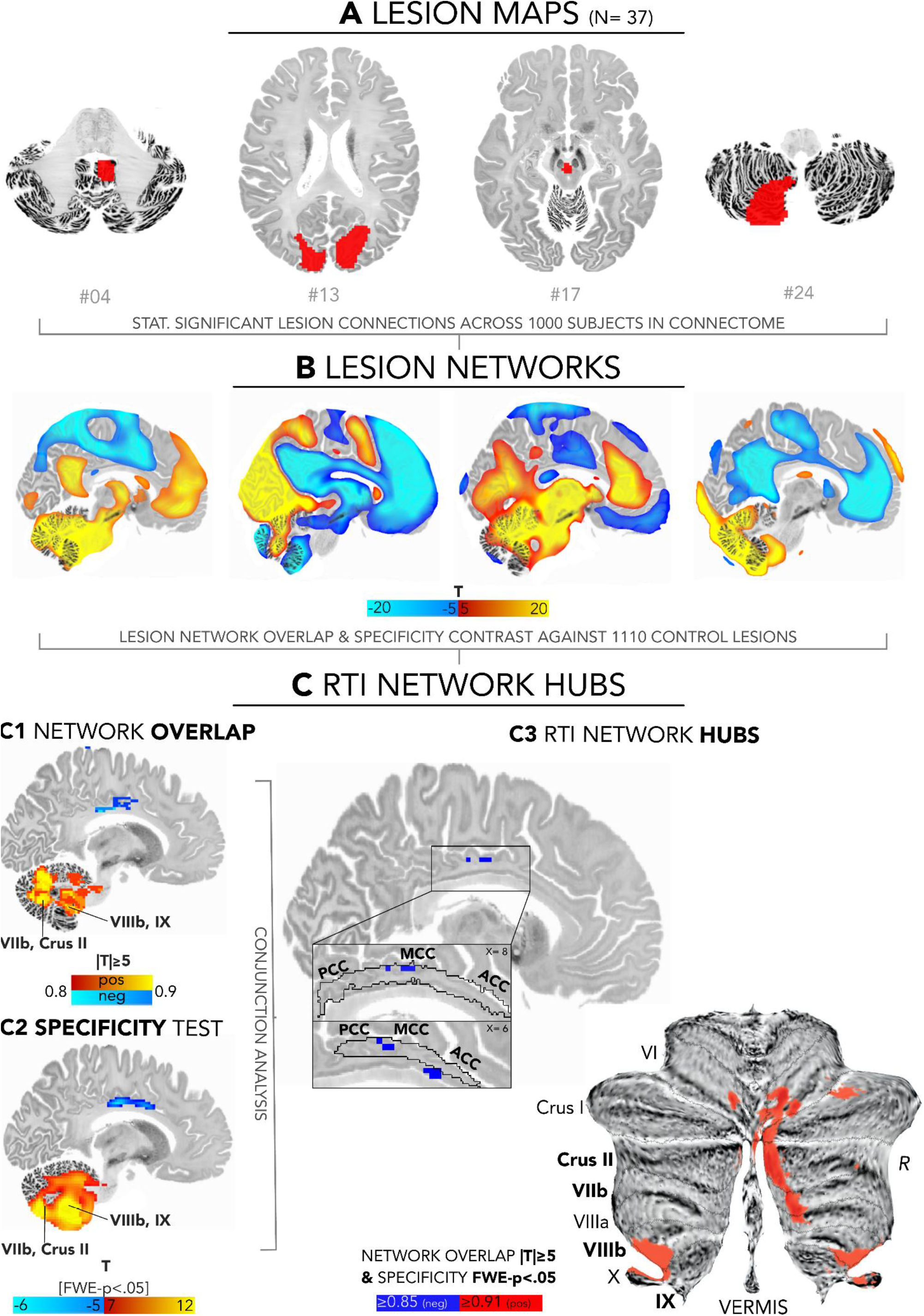
Lesions causing RTI map to common brain network hubs in the posterior cerebellum and cingulate cortex. A) Individual lesion locations associated with RTI (red) are mapped in a common reference space (MNI152). B) Functional connectivity between lesioned voxels and all other brain voxels is computed using the human functional connectome derived from 1,000 healthy subjects, resulting in a lesion network map for each lesion location. C1) Lesion network maps are thresholded (|T| ≥ 5, corresponding to FWE-corrected p < .05) and combined into an overlap map to identify regions consistently connected to RTI lesion locations, highlighting the posterior cerebellum and mid-cingulate cortex. C2) To identify RTI-specific connections, lesion network maps associated with RTI were compared with 1,110 lesions associated with 27 other neuropsychiatric symptoms or syndromes (voxel-wise FWE-corrected p < .05), resulting in a highly similar connectivity pattern centered on posterior cerebellar lobules VIIIb–IX, the paravermal zones of lobules VIIb– Crus II and the mid-cingulate cortex. C3) Combining the network overlap and specificity maps identified two principal RTI network hubs in the posterior cerebellum and mid-cingulate cortex that were both consistently and specifically connected to RTI lesion locations. Brain-wide peak regions of positive network overlap (≥91%) and specificity (peak T= 13) converged within 1 mm in posterior cerebellar lobules VIIIb–IX (MNI coordinates: −13, −53, −47 and 12, −51, −45), with secondary convergence in the right paravermal zone of lobule VIIb– Crus II (specificity peak T = 11; MNI coordinates: 14, −75, −43). Similarly, peak negative network overlap (≥89%) and specificity (peak T≤ −6.0) converged in the mid-cingulate cortex (MNI coordinates: 10, −16, 33). Positive connectivity is depicted in warm colors and red; negative connectivity (anticorrelation) in cool colors and blue.

In the posterior cerebellum, over 91% of lesions were significantly and positively connected to two anatomic-functional subregions: first, the (para-)vermal portions of lobules VI-VII and Crus I-II, and second, to the bilateral (peri-)floccular region encompassing lobules IX-X, and extending into lobule VIII. In the cingulate cortex, a peak of 89% of lesion locations was significantly connected to the mid-cingulate cortex (area A23c^51^) with neighboring areas occupying its border zones with the anterior and posterior cingulate cortex, respectively (Figure 3C3). Comparing RTI to 1,110 lesions associated with 27 other neuropsychiatric disorders and controlling for lesion volume, the posterior cerebellar-cingulate connectivity pattern remained highly specific to lesions associated with RTI (voxel-wise FWE-corrected p< .05, Figure 3C1-2). Notably, the results were robust to different assessors tracing the lesion locations (mean r 0.90 ± 0.16 across the 37 RTI lesions), different statistical thresholds (|T|= 5, 7, 9; Pearson r= 0.90 – 0.96) or connectome preprocessing pipelines^44^ (mean Pearson r= 0.94 ± 0.04), and remained similar when cases with cerebellar lesion location (18/ 37) were excluded (Pearson r= 0.88, Figure 4A). Exploratory permutation-based false-positive-rate analyses supported the thresholds used to define the RTI network hubs across lesion-network sensitivity, specificity, and their conjunction (Supplementary Figure 5).

**Figure 4.**
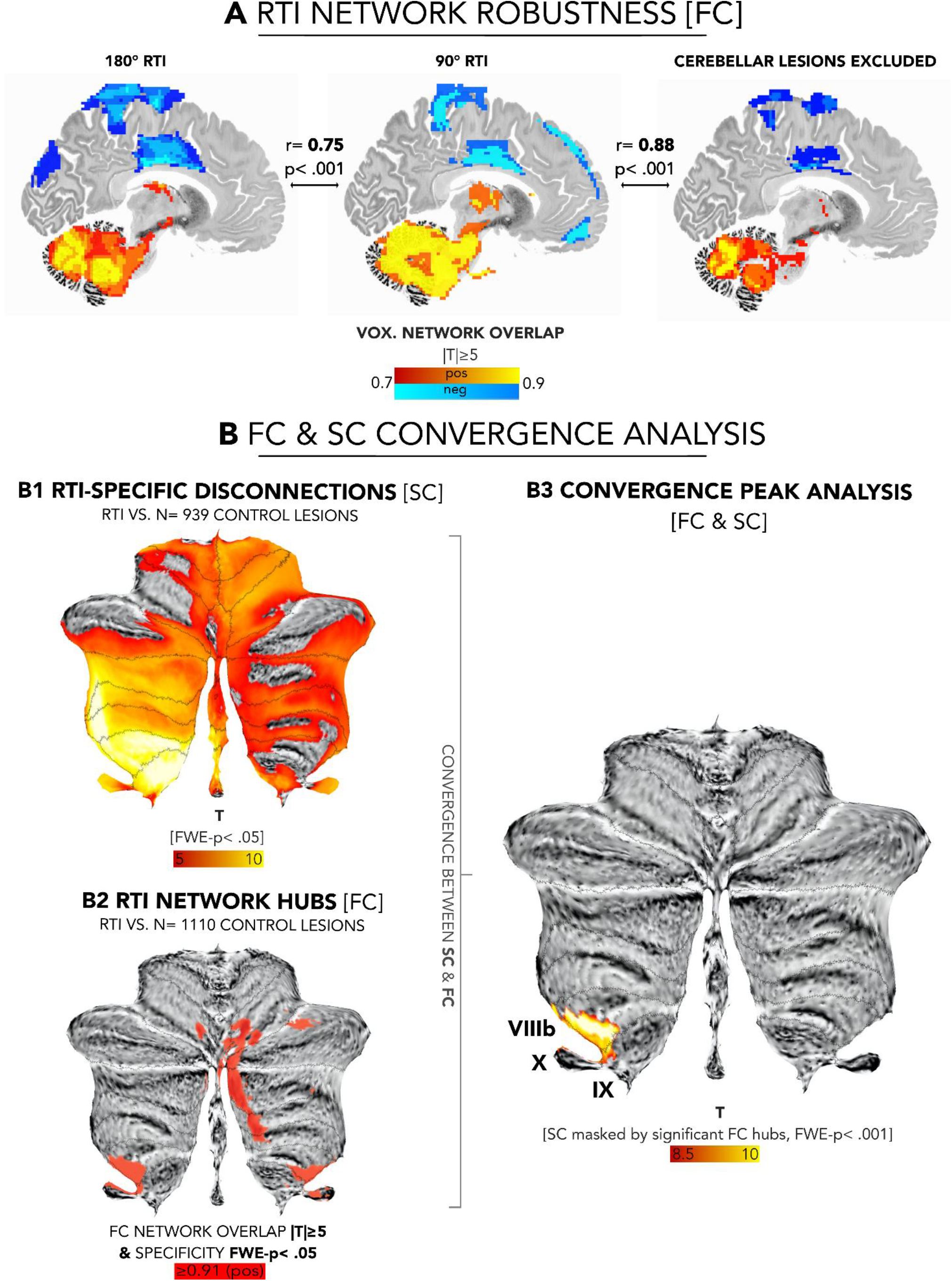
Functional network findings are independent of direct cerebellar lesion involvement and converge with structural lesion network mapping findings in the vestibulo-cerebellum. A. Functional network overlap maps derived separately from lesions causing 180° and 90° RTI showed similar spatial configurations (Pearson r = 0.75, p < .001). Importantly, repeating the analysis after excluding all cerebellar lesions (18/37) yielded a highly similar network map (Pearson r = 0.88, p < .001), indicating that cerebellar network convergence was not driven by lesions directly involving the cerebellum. B1. Structural disconnectome-based specificity analysis comparing lesion-induced disconnections associated with RTI (n= 1 versus those associated with other neuropsychiatric syndromes (n= 939) identifies RTI-specific disconnections within the posterior cerebellum (voxel-wise FWE-corrected p< .05). B2. Functional RTI network hubs derived from lesion network mapping, reproduced from Figure 3. B3. Overlay of structural and functional findings demonstrates peak convergence within the perifloccular portion of the RTI network hub, involving the cerebellar lobules IX, X and extending into VIII (FWE-corrected p < .001).

As a supplementary analysis, we used the two resulting RTI hub regions as seeds for another connectivity analysis aimed at identifying the brain voxels that are both positively connected to the positive cerebellar hubs and negatively connected to the negative mid-cingulate hub. Per definition, these voxels encompass a network that, when lesioned, can cause RTI. As expected, the resulting network encompasses 92% (34/37) of the neuroanatomically heterogeneous lesion locations causing RTI, primarily involving the occipito-parietal cortex, cerebellum, and brainstem (Supplementary Figure 6). Finally, we tested whether RTI subtypes show different network-level associations. However, no significant, subtype-specific patterns emerged in ROI-based label permutation similarity testing (p = 0.63) as well as in a whole-brain, voxel-wise two-sample t-test.

### Structural and functional networks linked to RTI converge on the posterior cerebellum

Given the established role of white matter pathways in verticality processing^9,28,61,62^ and to test whether our network findings generalize beyond a single connectivity modality, we repeated the analyses with structural network maps through a disconnectome analysis. This analysis again highlighted the posterior cerebellum, with peak disconnections converging onto the (peri-)floccular network hub identified in the prior functional connectivity analyses (voxel-wise FWE-corrected p< .001, Figure 4B1-3).

### RTI and milder disturbances of verticality perception dissociate at both the lesion and network level

Next, we aimed to determine whether RTI reflects an extreme phenomenology of the same neural substrates underlying milder disturbances of verticality perception, or whether it is associated with a distinct network architecture. Importantly, this comparison also provided a granular phenotype-matched test of network specificity, in line with recent methodological arguments favoring clinically relevant control lesions for specificity testing in lesion network mapping^35,63^.

To this end, we compared RTI to a cohort with lesions causing canonical signs of disturbed verticality perception, including ocular tilt reaction and/or SVV tilts^19,20,33^. Across 185 cases assembled from two independent lesion datasets (mean SVV tilt: 7.96 ± 5.85°, maximum 30°), lesions causing milder verticality disturbances similarly showed substantial anatomical heterogeneity with little spatial overlap (maximum overlap: 21/185 cases, 11%). Lesion overlap was located in the right posterior insula, a region that was also highlighted in an exploratory VLSM analysis (FWE-corrected p< .05, Supplementary Figure 7A-B). Importantly, the two cohorts were comparable with respect to the distribution of cortical versus subcortical lesion distribution (Fisher’s exact test p = 0.31).

Extending this approach to network-level patterns, we next applied the same lesion network mapping framework we used for RTI lesions to the set of lesions associated with milder disturbances of verticality perception. While the resulting lesion networks converged on hub regions mainly in the intermediate cerebellar lobules VI–VIII, their topography was notably distinct from that observed in RTI, with minimal overlap in the paravermal portion of lobule VI (Supplementary Figure 8).

Directly comparing lesion connectivity between RTI and milder SVV tilts through a whole-brain two-sample t-test revealed significant connectivity differences in both the cingulate cortex and posterior cerebellum (voxel-wise FWE-corrected p< .05; Figure 5). Secondary clusters of significance were identified in the bilateral posterior insulae (more negatively connected in RTI) and lateral occipital cortex (more positively connected in RTI; Supplementary Figure 8). Disconnectome-based comparisons yielded sparse differences surviving correction for multiple comparisons. Exploratory inspection of the full, unthresholded maps however showed relatively greater involvement of occipito-temporo-insular pathways in RTI and greater involvement of pontine and midbrain tegmental pathways in milder verticality disturbances (Supplementary Figure 7C).

**Figure 5.**
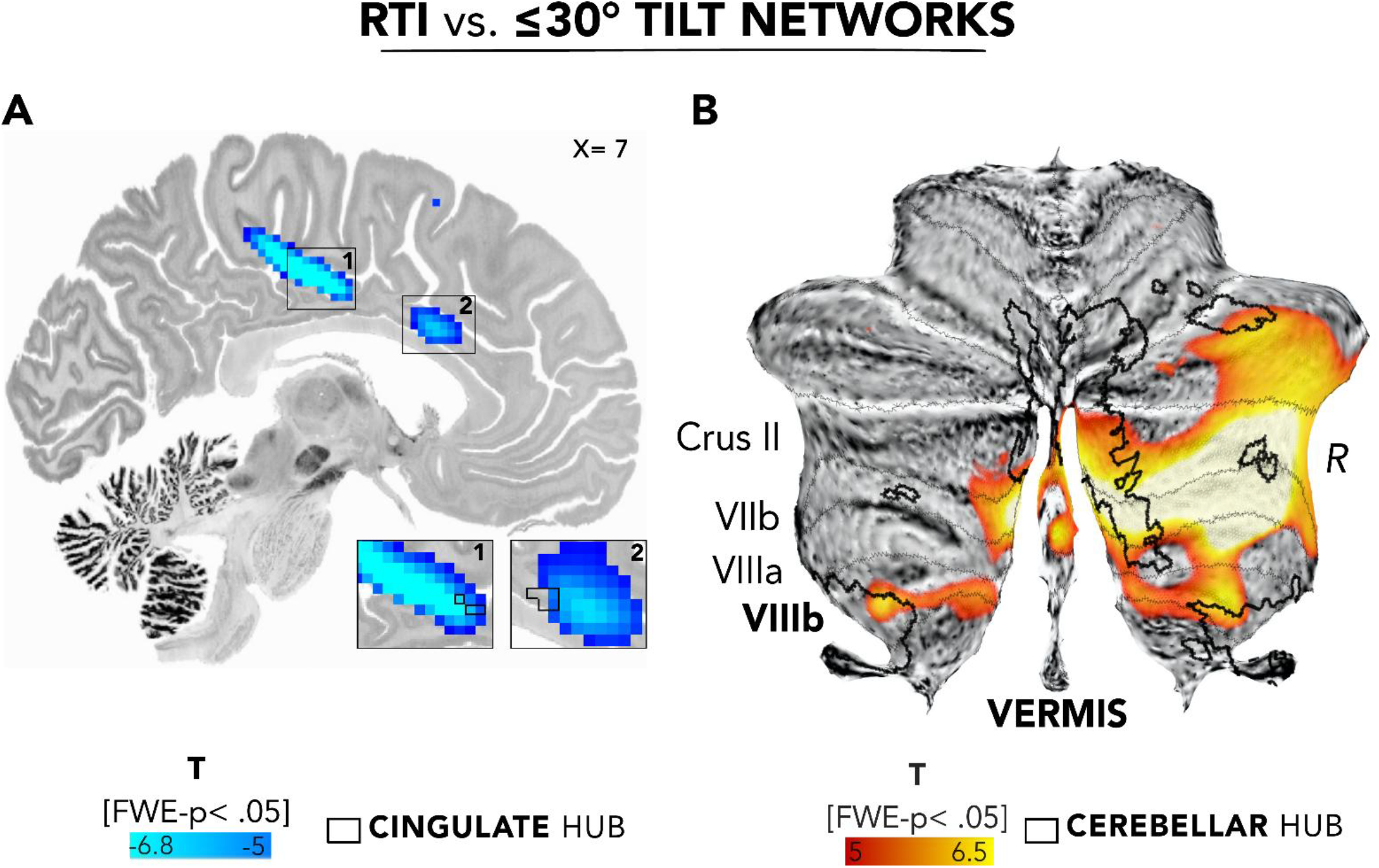
Extreme and mild disturbances of verticality perception are associated with distinct brain networks. Direct comparison of lesion network maps associated with RTI and milder disturbances of verticality perception (≤30° tilt) using a voxel-wise 2-sample t-test. A. RTI is associated with significantly stronger (negative) connectivity to the mid-cingulate cortex compared with lesions causing milder tilt deviations (voxel-wise FWE-corrected p< .05), encompassing the previously identified RTI hub region (black outlines). Insets 1 & 2 show a magnification of the relationship of the connectivity peaks and the topography of the lesion-derived RTI hub in the cingulate. B. RTI lesions are also associated with significantly stronger connectivity to posterior cerebellar regions involving the (para-)vermal portions of posterior lobules Crus II, VIIb–VIIIb (voxel-wise FWE-corrected p< .05), again encompassing previously identified RTI hub regions.

### Direct electrical stimulation sites causing vestibular sensations are preferentially connected to RTI network hubs

Current pathophysiological models of RTI invoke dysfunctional interaction of higher-order vestibular and visual networks^16,24,31,64^. Because our lesion network analyses revealed substantial differences between RTI and milder disturbances of verticality perception - as established disorders of lower-to-intermediate-level vestibular processing^4^ - we next asked whether the lesion-derived RTI hubs are preferentially embedded within vestibular cortical networks.

To test this directly, we leveraged an independent retrospective dataset of DES sites associated with vestibular sensations (including vertigo, dizziness and self-motion illusions; n = 57) and non-vestibular effects or no perceptual effects (n = 452). Using the lesion-derived RTI hubs as independently defined ROIs, label permutation testing showed that vestibular DES sites were significantly more strongly connected to both RTI hubs than control stimulation sites (10,000 permutations; cingulate hub: p < .001, Hedges’ g = 0.62; posterior cerebellar hub: p = .014, Hedges’ g = 0.34; Figure 6A-B). As a supplementary whole-brain analysis, we treated DES foci analogously to lesions within the network mapping framework, confirming specific connectivity in the mid-cingulate cortex, encompassing the RTI hub region (voxel-wise FWE-corrected p < .05, Supplementary Figure 9).

**Figure 6.**
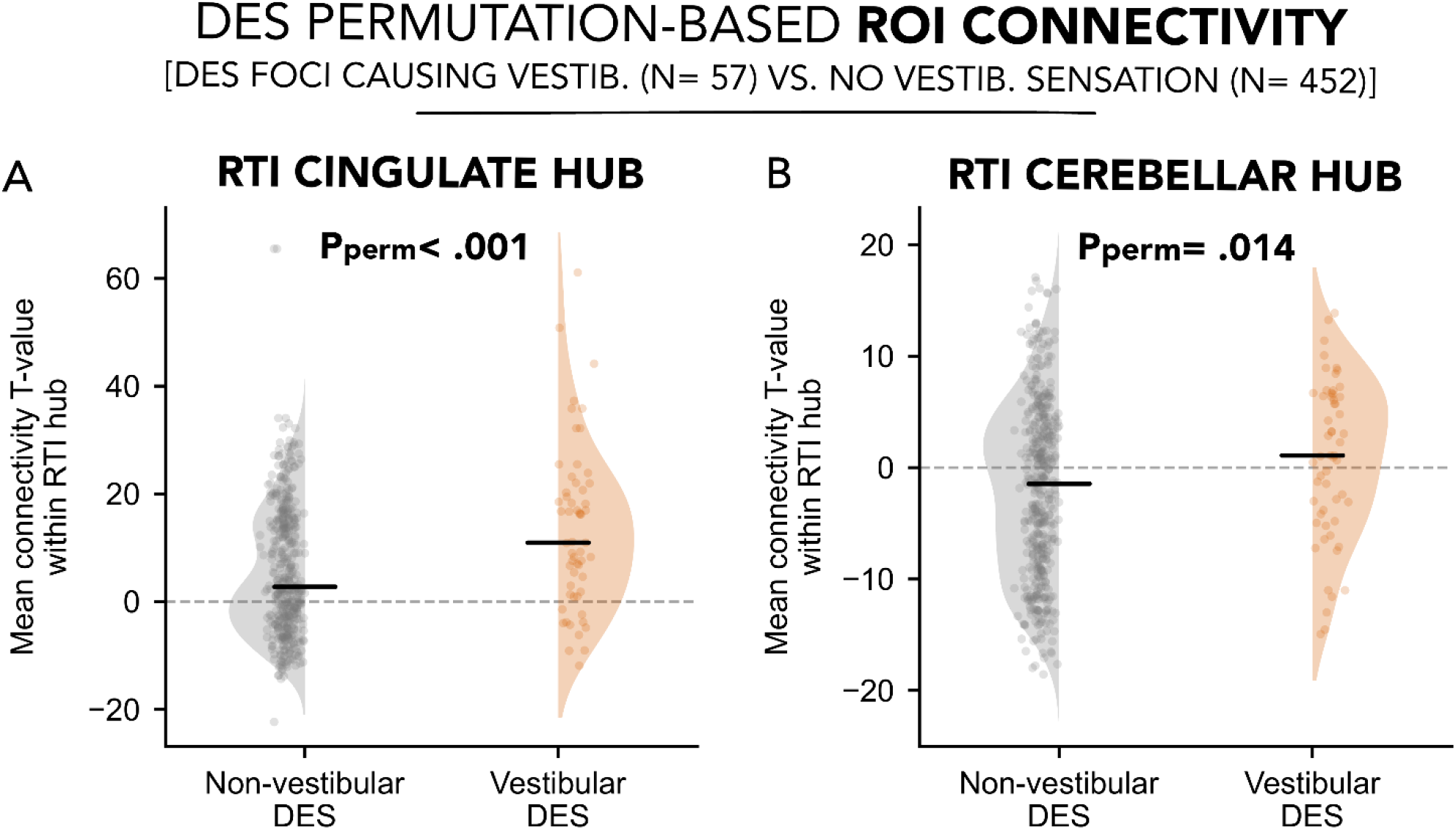
Vestibular stimulation sites preferentially connect to RTI network hubs. ROI-based permutation analyses within the lesion-derived RTI hubs in the mid-cingulate cortex (A) and posterior cerebellum (B). Vestibular DES sites showed significantly stronger connectivity to both RTI hubs than non-vestibular DES sites (10,000 permutations; cingulate hub: p< .001, Hedges’ g= 0.62; cerebellar hub: p = .014, Hedges’ g= 0.34). Each point represents one DES focus network; horizontal bars denote group medians.

## DISCUSSION

Reliable perception of verticality is fundamental for spatial orientation and postural stability, and its disruption contributes to falls and disability across neurological disorders or during aviation and space flight. Specifically, RTI is a transient disorder in which the orientation of the visual environment is perceived as significantly tilted relative to gravity, most often quantized at 90° or 180°^65^. In this regard, RTI has been proposed as a model disorder to probe the multisensory computations underlying verticality perception, but its neural basis has remained unresolved, in part because causative lesions can be anatomically scattered throughout cortical as well as subcortical brain regions.

In 37 cases of lesion-induced RTI, we found clinical and anatomical complementary support for this network architecture: structural connectivity converged on the posterior cerebellar hub, while independent DES data revealed preferential connectivity of vestibular stimulation sites to both lesion-derived RTI hubs. Together, these findings suggest a two-fold circuit-level substrate for RTI.

The anatomical localization of the hubs identified by our multimodal brain mapping approach provides context for this interpretation. The posterior cerebellar hub – functionally connected to 91% of RTI lesions – chiefly occupies two vestibulo-cerebellar territories: a (para-)vermal region, linked to internal models of spatial orientation and self-motion relative to gravity^66,67^, and a (peri-)floccular region, involved in integrating vestibular and visual signals for gaze stabilization in three-dimensional (3D) space^68^. Notably, structural disconnectome mapping independently environment, closely resembling the RTI phenomenology^72–75^. Similarly, RTI can be induced by vestibular stimulation in patients with brainstem lesions only in illuminated conditions^31^, while visual information has been shown to dominate verticality estimates in people with cerebellar degeneration^70^.

Within this framework, we hypothesize that in lesion-induced RTI, disconnection of posterior vestibulo-cerebellar circuitry may degrade the reliability of vestibular verticality estimates, leading to a shift of perceptual inference toward visual scene information. But why should such a shift produce the discrete, cardinal reorientations characteristic of RTI rather than a graded perceptual tilt? Environmental structure may provide one explanation. Natural and particularly built environments contain strong cardinal cues such as the horizon, walls or ceilings, and these cardinal orientations are preferentially encoded over oblique orientations in the visual cortex^76,77^. Together, these external and internal constraints could bias perceptual inference toward a limited set of alternative but internally coherent interpretations of the visual world during RTI. Consistent with this hypothesis, eye closure has been reported to abolish RTI in some cases^16^, and among our cases with available contextual information, episodes occurred almost exclusively indoors (20/21), where rectilinear environmental cues provide particularly strong cardinal references for upright. The cerebellar association alone, however, appears insufficient to explain RTI. The majority of RTI lesions (19/37) were located outside the cerebellum while converging on the same network. More generally, RTI remains rare relative to the frequency of cerebellar strokes^16,78^. Moreover, lesions to closely neighboring cerebellar sites linked to tremor^79^, cervical dystonia^80^ or cognitive-affective syndromes^81^ are not typically associated with RTI. Together, these observations suggest a potentially two-fold network architecture.

Indeed, a second RTI-specific hub was found in the mid-cingulate cortex. This hub, chiefly corresponding to area A23c^51^, was negatively connected in up to 89% of RTI lesions and independently supported by causal evidence from DES. Importantly, this cingulate territory was one of the two regions most strongly distinguishing RTI from phenotypically related, milder disturbances of verticality perception, suggesting that on a network level, RTI cannot simply be understood as an extreme form of graded verticality perception errors. Converging evidence from our stimulation mapping analyses and previous neurophysiological studies implicates the cingulate cortex in two multisensory computations: higher-order vestibular processing and representation of one’s own body and self-location^10,82–89^. More specifically, large-scale studies showed that DES to the mid-cingulate cortex (including A23c) most frequently induces body-anchored and gravity-related sensations, such as a sense of weightlessness or floating of single body parts or the body as a whole^86,89–91^. Based on this evidence, we hypothesize that the cingulate hub may contribute to anchoring gravity-referenced information within a coherent representation of self-orientation relative to the external world^92^. Therefore, we propose a tentative circuit-level model in which the posterior cerebellar hub may contribute to multisensory estimation of verticality, whereas the cingulate hub may relate this estimate to a body-centered representation of self-orientation within space. From this perspective, RTI may emerge when a specific disruption of this cerebello-cingulate network destabilizes the gravity-referenced relationship between body and external space to allow a dramatically erroneous estimate of spatial orientation to become transiently stabilized^10,90^.

Notably, canonical cortical vestibular regions such as the posterior insula did not emerge as principal RTI hubs. Although the posterior insula was associated with milder verticality disturbances and also showed weaker effects in functional and structural specificity analyses with respect to RTI, these were substantially less pronounced than the posterior cerebellar and cingulate findings. Thus, while RTI appears to involve regions embedded within the broader vestibular network^10,93^, its network anatomy differs from that associated with vertigo and graded verticality errors, which have been linked more strongly to posterior insular circuitry^25^. This dissociation may therefore provide an anatomical framework for the concept of distinct “higher vestibular functions” as proposed by Brandt and colleagues^23^.

Several limitations warrant emphasis. First, our data are primarily drawn from published case reports, which are subject to ascertainment and publication bias. We aimed to mitigate this by extracting detailed clinical characteristics beyond the presence of RTI, which grossly aligned with results from a published systematic review. Second, lesions were traced from 2D figures of variable quality, which might have affected accuracy. We mitigated this by following best practice standards for lesion segmentation^38^, and also by quantifying inter-assessor reliability, which was high (mean r = 0.90). Third, lesion network mapping relied on a normative functional connectome rather than patient-specific connectivity and has recently been the subject of methodological debate regarding reproducibility and specificity^63^. We therefore deliberately combined multiple connectivity methods, independent datasets, recent validated analytical pipelines, and permutation-based assessment of false-positive rates to test the robustness of the thresholds used to define the RTI network^35,50,94^. Fourth, the mid-cingulate hub was defined by negative connectivity, whose physiological meaning remains debated, as anticorrelations may reflect inhibitory network interactions but are also influenced by global signal regression^43^. Nevertheless, its robustness across preprocessing pipelines and convergence with vestibular DES support its biological relevance.

In conclusion, beyond investigating a rare neurological syndrome, RTI provides a lesion-based model for localizing the neural circuitry supporting verticality perception. By identifying a convergent and specific cerebello-cingulate network, our findings provide an anatomical framework for the multisensory computations thought to underlie verticality perception and body-anchored self-orientation. By localizing this circuitry, our findings generate testable hypotheses for experimentally probing and modulating human verticality perception and the underlying multisensory computations, with potential translational applications^8,13,95,96^.

## Data Availability

Lesion masks in MNI space are available upon request to the corresponding author. The code for lesion connectivity analysis is freely distributed via the Lead-DBS toolbox (www.lead-dbs.org). The fully preprocessed version of the human resting-state fMRI connectome is publicly available on Harvard Dataverse (https://dataverse.harvard.edu/dataset.xhtml?persistentId=doi:10.7910/DVN/ILXIKS) and https://zenodo.org/records/4905738.

https://zenodo.org/records/4905738

## Acknowledgements

The authors thank the scientific community involved in publishing the case studies underlying this work.

