## Supplementary materials for "Lesions causing room tilt illusion link a cerebello-cingulate brain network to verticality perception"

**Extended case descriptions and clinical characteristics**

Lesions causing milder disturbances of verticality perception

For comparisons between lesion locations and brain networks across tilt severities, a dataset of 185 lesions associated with milder disturbances of verticality perception (≤30° tilt deviation) was assembled, as outlined in the main methods.

This dataset comprised 89 cases identified through a PRISMA-based literature review focusing on lesion-induced milder misperceptions of verticality operationalized as ocular tilt reaction (OTR) and/ or documented deviations of the subjective visual vertical (SVV). Mean age was 54.5 ± 16.3 years (range 5–87 years), 61% were male. Lesions were predominantly infratentorial (82/89, 92%), whereas 7 lesions involved supratentorial structures. Assessment of verticality perception beyond bedside examination was documented in 36 cases, with SVV angles available in 29 cases (mean absolute SVV tilt 10.4 ± 7.1°, range 3 – 30°). Detailed demographic and lesion-level characteristics are provided in Supplementary Table 2.

In addition, a local multi-institutional convenience sample comprising 96 lesion cases associated with pathological deviations of the SVV (≥2.5°, assessed using paradigms such as the visual dome or bucket test^1^) was included (mean absolute SVV: 6.00 ± 3.86°, range 2.5 – 24.82°). Within this cohort, 57 lesions were located cortically and 39 involved subcortical regions, with a majority located on the right hemisphere (94%). Detailed demographic and clinical characteristics of this cohort have been reported previously^2–4^.

**Supplementary Tables**

| **ID** | **DOI** | **case_n_theirs** | **Tilt_degree** |
| --- | --- | --- | --- |
| RTI01 | 10.1007/s00415-012-6536-0 | 1 | 180 |
| RTI02 | 10.1007/s00415-012-6536-0 | 2 | 90 |
| RTI03 | 10.1007/s00415-012-6536-0 | 3 | 90 |
| RTI04 | 10.1007/s00415-012-6536-0 | 4 | 180 |
| RTI05 | 10.1007/s00415-012-6536-0 | 9 | 180 |
| RTI06 | 10.1016/S0010-9452(88)80044-3 | 1 | 180 |
| RTI07 | 10.1136/jnnp.55.3.188 | 1 | 180 |
| RTI08 | 10.1007/s00415-006-0124-0 | 1 | 180 |
| RTI09 | 10.1007/s10072-019-04041-9 | 1 | 90 |
| RTI10 | 10.1136/jnnp.56.4.423 | 1 | 180 |
| RTI11 | 10.1007/s004150050123 | 1 | 180 |
| RTI12 | 10.1001/archneur.55.10.1362 | 1 | 180 |
| RTI13 | 10.1001/archneur.55.10.1362 | 2 | 180 |
| RTI14 | 10.1016/S0140-6736(06)68544-9 | 1 | 180 |
| RTI15 | 10.1007/s10072-011-0781-3 | 1 | 180 |
| RTI16 | 10.12622/2317-3076jhbs.v2i4.97p.224.2014 | 1 | 180 |
| RTI17 | archneur.1996.00550120071018 | 1 | 90 |
| RTI18 | archneur.1996.00550120071018 | 2 | 90 |
| RTI19 | archneur.1996.00550120071018 | 3 | 90 |
| RTI20 | 10.1159/000083272 | 1 | 90 |
| RTI21 | 10.1097/WNO.0b013e31815bfc02 | 1 | 90 |
| RTI22 | Local case 1 |  | 90 |
| RTI23 | 10.4103/0028-3886.100709 | 1 | 90 |
| RTI24 | 10.1007/s00115-013-3965-7 | 1 | 90 |
| RTI25 | 10.1136/jnnp.55.3.188 | 1 | 180 |
| RTI26 | 10.1016/j.yebeh.2005.08.012 | 1 | 180 |
| RTI27 | 10.1177/1352458507082045 | 1 | 180 |
| RTI28 | Local case 2 |  | 90 |
| RTI29 | 10.1016/s0028-3843(14)60158-4 | 1 | 90 |
| RTI30 | 10.1097/WNO.0000000000001501 | 1 | 180 |
| RTI31 | 10.1016/j.neuarg.2012.11.004 | 5 | 180 |
| RTI32 | 10.1097/00041327-200306000-00008 | 1 | 45 |
| RTI33 | 10.33588/rn.4612.2008121 | 1 | 180 |
| RTI34 | 10.1080/01658100801951790 | 1 | 180 |
| RTI35 | Local case 3 |  | 180 |
| RTI36 | 10.1136/bcr-2022-253965 | 1 | 90 |
| RTI37 | Local case 4 |  | 180 |

Supplementary Table 1. **Case list and references of RTI lesions.**

| **PMID** | **Sex** | **Lesion side** | **Lesion location** | **OTR** | **SVV tilt** | **Tilt direction** | **Abs. SVV tilt (°)** |
| --- | --- | --- | --- | --- | --- | --- | --- |
| 20379734 | F | L | Insula | NA | Y | L | NA |
| 32759590 | M | L | Temporal | N | Y | R | NA |
| 12588938 | F | L | Temporal | N | Y | R | 6 |
| 8154866 | F | R | Insula, parietal | N | Y | L | 6 |
| 15156094 | M | L | Temporal | N | Y | L | 5 |
| 36633670 | F | R | Parietal | NA | Y | L | NA |
| 24581670 | F | R | Parietal, occipital | N | Y | R | 3.2 |
| 34966124 | M | R | Occipital, thalamus | Y | NA | NA | NA |
| 24249795 | F | L | Midbrain, thalamus | Y | NA | NA | NA |
| 25562880 | M | L | Cerebellum | Y | NA | NA | NA |
| 9629842 | M | L | Midbrain, thalamus | Y | NA | NA | NA |
| 9629842 | M | R | Midbrain, thalamus | Y | NA | NA | NA |
| 9629842 | F | R | Midbrain, thalamus | Y | NA | NA | NA |
| 22170943 | M | R | Pons | Y | NA | NA | NA |
| 26928363 | M | B | Cerebellum, midbrain | Y | NA | NA | NA |
| 19909722 | F | R | Midbrain, thalamus | Y | NA | NA | NA |
| 36067407 | M | L | Cerebellum | Y | Y | R | 6 |
| 14610132 | M | L | Cerebellum, pons | NA | Y | R | 12 |
| 22346205 | M | L | Pons, medulla | Y | NA | NA | NA |
| 16599282 | F | B | Thalamus | Y | NA | NA | NA |
| 25025301 | F | R | Pons | Y | Y | L | 5 |
| 25692514 | F | R | Medulla | Y | Y | L | 3.5 |
| 19144523 | F | R | Pons | Y | NA | NA | NA |
| 37497260 | M | R | Midbrain, thalamus | Y | NA | NA | NA |
| 15826990 | M | L | Medulla | Y | Y | L | 5 |
| 15826990 | M | R | Medulla, cerebellum | Y | Y | R | 5 |
| 31736578 | F | R | Medulla | Y | NA | NA | NA |
| 33538908 | M | R | Cerebellum, medulla | NA | NA | NA | NA |
| 33538908 | M | R | Pons | Y | NA | NA | NA |
| 22177934 | M | B | Midbrain, cerebellum | Y | NA | NA | NA |
| 7943131 | F | R | Pons, cerebellum | Y | NA | NA | NA |
| 7943131 | M | R | Pons, cerebellum | Y | Y | R | NA |
| 7943131 | F | L | Pons | Y | NA | NA | NA |
| 7943131 | M | R | Medulla | Y | NA | NA | NA |
| 27448548 | M | L | Pons | Y | NA | NA | NA |
| 2215939 | M | R | Midbrain, thalamus | Y | Y | L | 17 |
| 2215939 | M | R | Midbrain, thalamus | Y | Y | L | 20 |
| 2215939 | F | R | Midbrain, thalamus | Y | NA | NA | NA |
| 2215939 | M | R | Thalamus | Y | NA | NA | NA |
| 7103431 | M | L | Midbrain | Y | NA | NA | NA |
| 34974486 | M | L | Pons, medulla | Y | NA | NA | NA |
| 1734314 | M | R | Pons, medulla | Y | NA | NA | NA |
| 25882476 | M | L | Medulla | N | Y | L | 18 |
| 25882476 | M | R | Medulla | N | Y | R | 13.5 |
| 27647583 | M | R | Midbrain, thalamus | Y | NA | NA | NA |
| 21769260 | F | L | Thalamus | Y | Y | R | NA |
| 21769260 | F | L | Thalamus | NA | Y | L | NA |
| 16291911 | M | L | Pons | Y | Y | L | 17 |
| 19577773 | F | L | Pons | NA | Y | L | 7.5 |
| 33486342 | M | L | Pons | Y | NA | NA | NA |
| 36814996 | F | L | Cerebellum | Y | NA | NA | NA |
| 33599814 | NA | R | Pons | NA | Y | L | 17.2 |
| 33599814 | NA | L | Pons | NA | Y | R | 10.9 |
| 33599814 | NA | L | Pons | NA | Y | R | 3 |
| 33599814 | NA | R | Pons | NA | Y | L | 9.8 |
| 24925102 | F | L | Midbrain, thalamus | Y | NA | NA | NA |
| 25432932 | F | R | Midbrain, thalamus | Y | Y | L | NA |
| 19145125 | M | L | Thalamus | Y | NA | NA | NA |
| 3228274 | F | R | Medulla | NA | NA | NA | NA |
| 9270583 | M | L | Cerebellum | NA | Y | R | 7.7 |
| 8410014 | M | L | Medulla | Y | NA | NA | NA |
| 29434137 | M | L | Pons | Y | NA | NA | NA |
| 16127261 | M | B | Midbrain, thalamus | Y | NA | NA | NA |
| 19038407 | M | R | Thalamus | NA | NA | NA | NA |
| 19038407 | F | R | Thalamus, basal ganglia, midbrain, frontal | Y | NA | NA | NA |
| 9532539 | M | R | Midbrain | Y | NA | NA | NA |
| 19952901 | M | R | Midbrain | Y | NA | NA | NA |
| 23084391 | M | L | Midbrain | Y | Y | R | 5 |
| 23370610 | F | R | Cerebellum | N | Y | L | 7.4 |
| 24899334 | M | L | Midbrain, thalamus | Y | NA | NA | NA |
| 12566042 | F | R | Pons, midbrain | Y | NA | NA | NA |
| 22142667 | M | L | Medulla | Y | Y | L | 3 |
| 19365047 | F | L | Thalamus | Y | NA | NA | NA |
| 21786085 | M | L | Cerebellum | Y | Y | R | 16 |
| 35667839 | F | L | Pons, medulla | NA | NA | NA | NA |
| 25515599 | M | R | Cerebellum | N | Y | L | 22.5 |
| 33252541 | M | L | Pons | Y | NA | NA | NA |
| 8970457 | F | B | Midbrain | Y | Y | R | 3 |
| 8970457 | M | R | Medulla | Y | Y | R | 30 |
| 8970457 | F | L | Pons | Y | N | NA | NA |
| 11096232 | M | R | Midbrain | Y | NA | NA | NA |
| 11096232 | F | R | Pons | Y | NA | NA | NA |
| 22001484 | M | L | Pons | Y | NA | NA | NA |
| 23037477 | M | B | Cerebellum | NA | NA | NA | NA |
| 25274240 | M | L | Cerebellum | Y | NA | NA | NA |
| 16087906 | M | L | Cerebellum | Y | NA | NA | NA |
| 16087906 | F | L | Cerebellum | Y | NA | NA | NA |
| 16087906 | F | B | Cerebellum | Y | NA | NA | NA |
| 28844105 | M | R | Cerebellum | Y | Y | L | 17 |

Supplementary Table 2. **Demographic and clinical characteristics of literature-derived cases with milder verticality misperceptions identified through a PRISMA-based review**. SVV direction and absolute SVV tilt are reported where available. Absolute SVV tilt denotes the magnitude of deviation from true vertical irrespective of direction. Abbreviations: B, bilateral; L, left; NA, not available; R, right; OTR, ocular tilt reaction; SVV, subjective visual vertical.

**Supplementary Figures**

**Supplementary Figure 1**. PRISMA search workflow for lesions causing RTI.

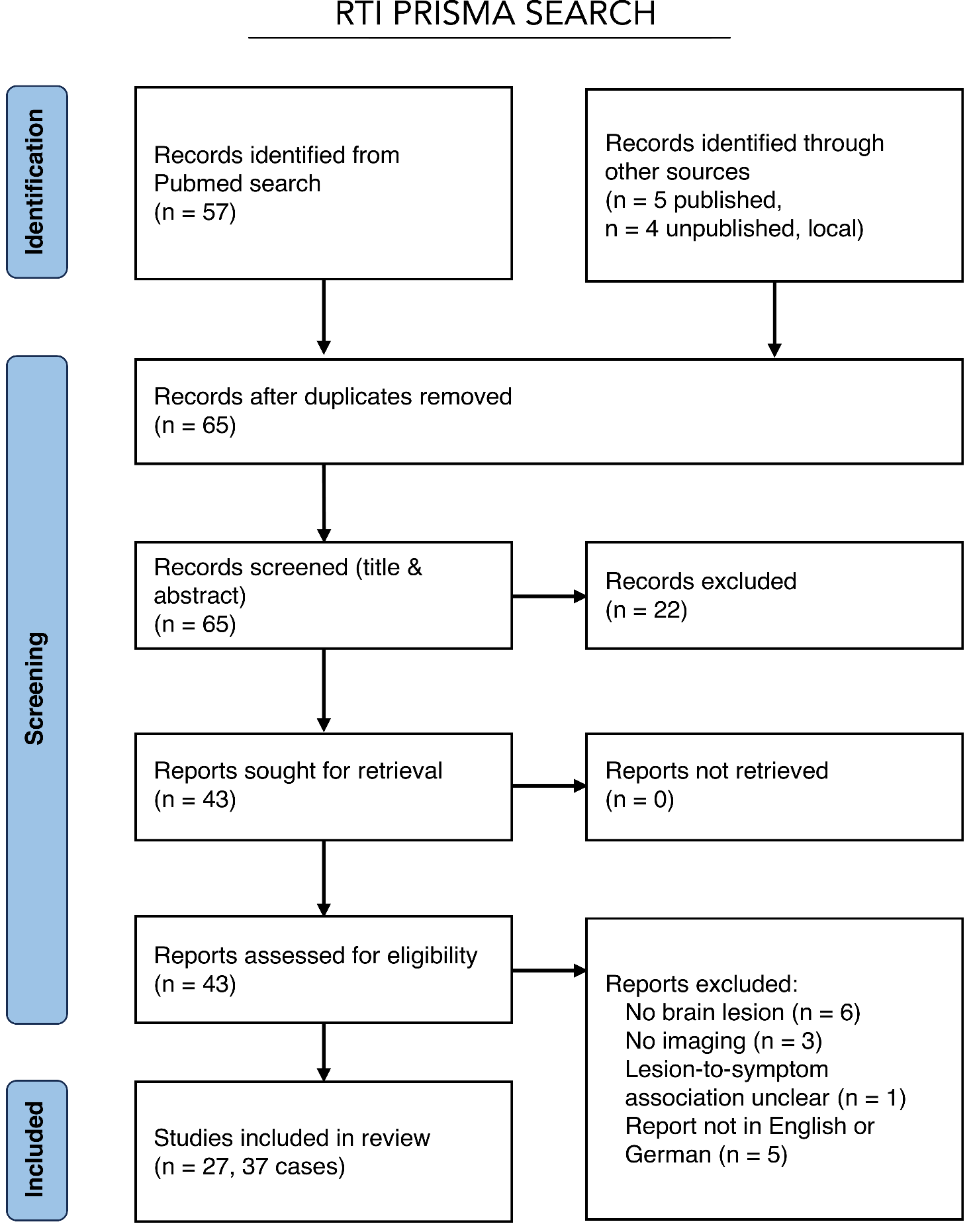

**Supplementary Figure 2**. PRISMA search workflow for lesions causing milder disturbances of verticality perception.

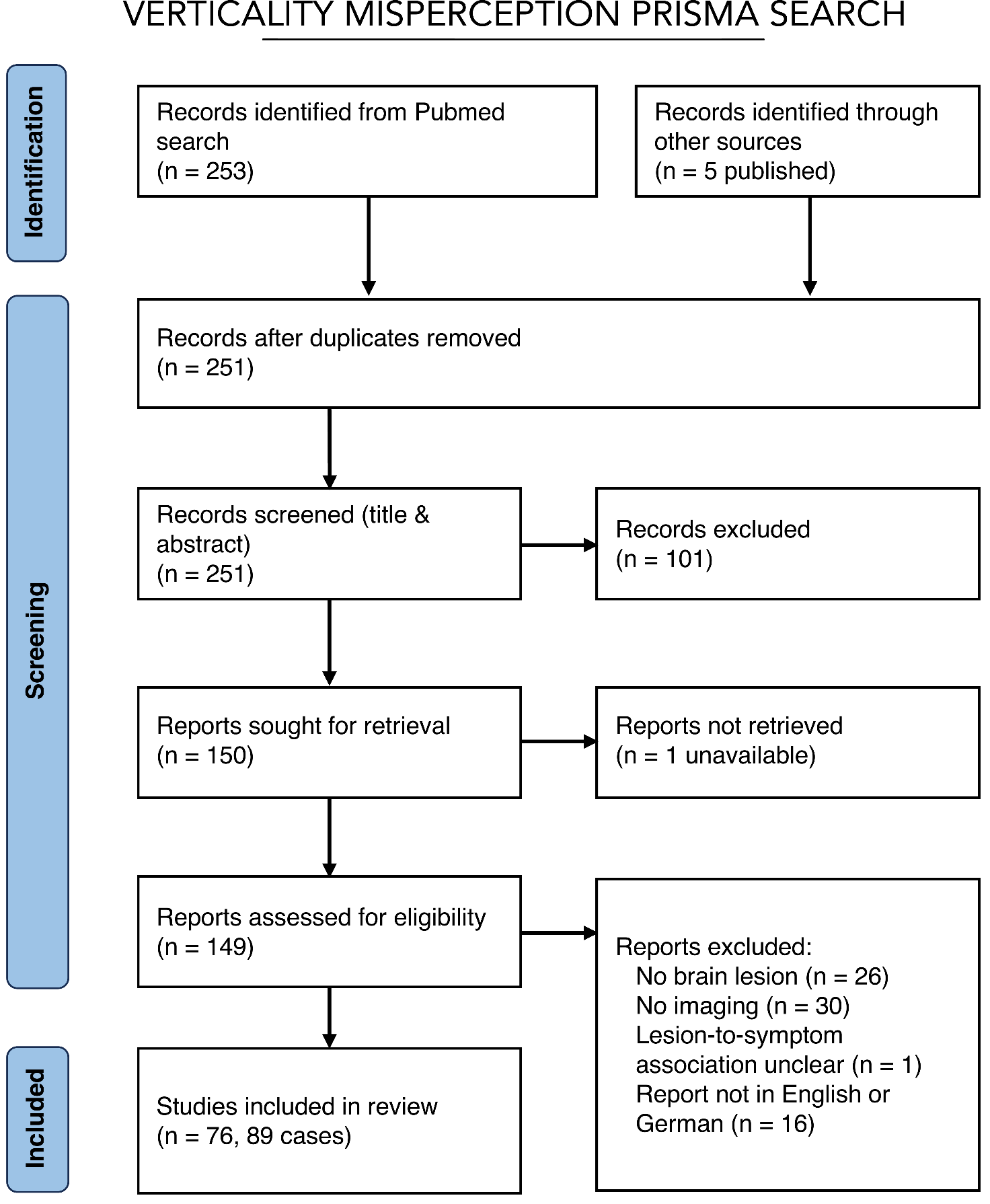

**Supplementary Figure 3**. RTI lesion masks.

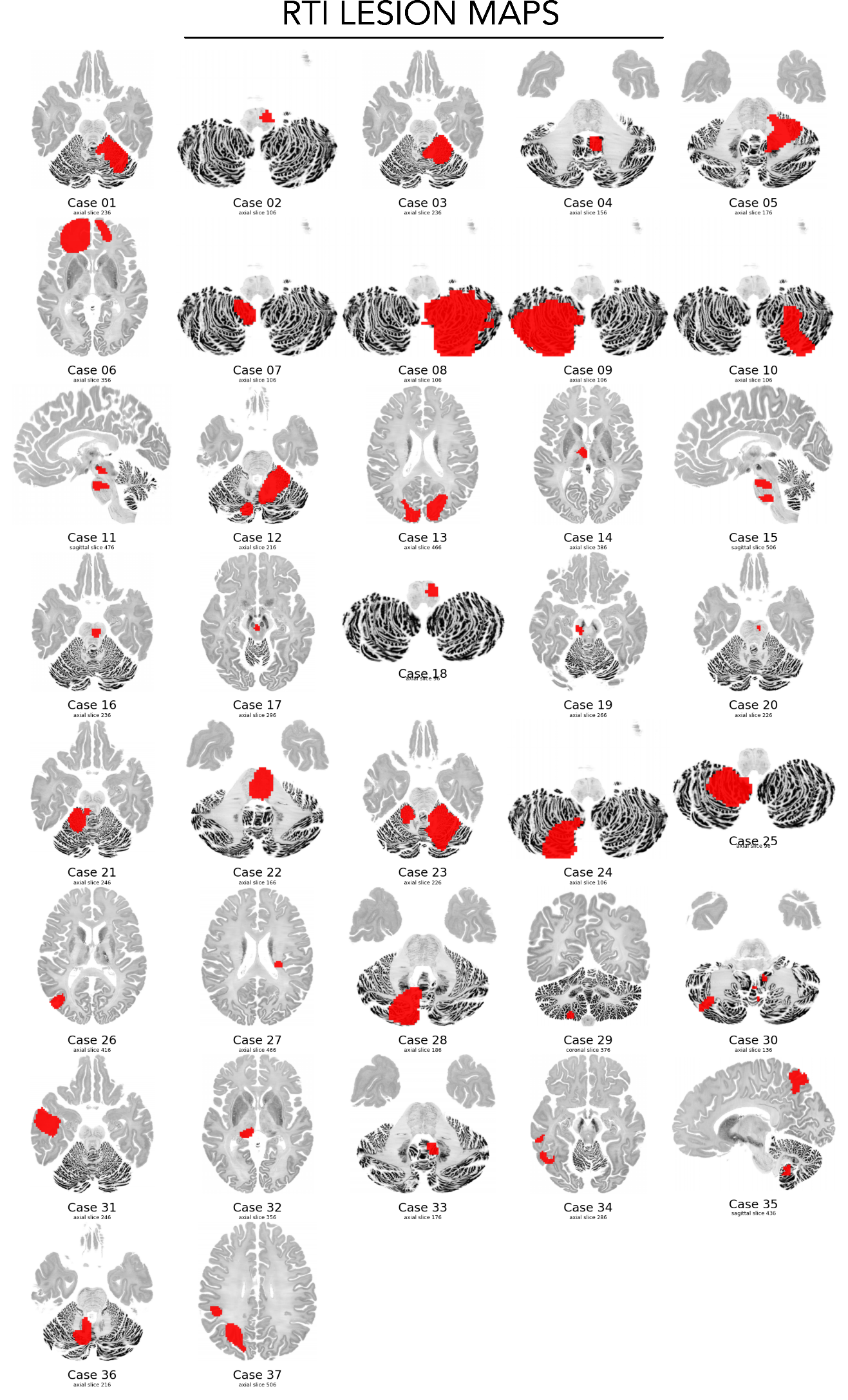

**Supplementary Figure 4**. RTI lesions overlap analysis.

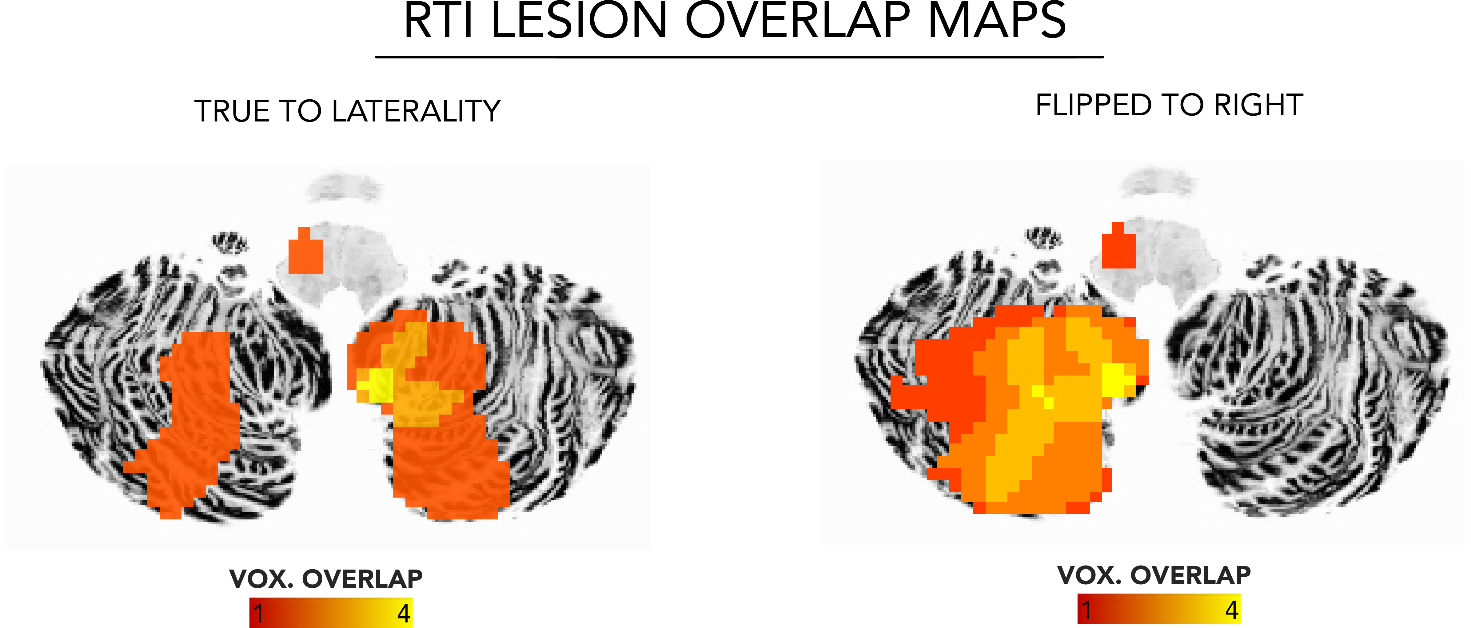

**Supplementary Figure 5.** Permutation-based false-positive-rate analysis of RTI lesion network mapping thresholds. A. Empirical false-positive rates for lesion network overlap across combinations of individual lesion-network T-map thresholds (|T| = 5, 7, 9, and 11) and group-level lesion network overlap thresholds. Increasing either threshold progressively reduced the proportion of false-positive findings under permutation. The red dashed line indicates an empirical false-positive rate of 5% (α = 0.05). B. Empirical false-positive rates for the conjunction of lesion network overlap and RTI specificity across different overlap and specificity T-statistic thresholds. The threshold combinations used to define the RTI network hubs were associated with false-positive rates below 1% (see Main Figure 3). The red dashed line indicates α = 0.05.

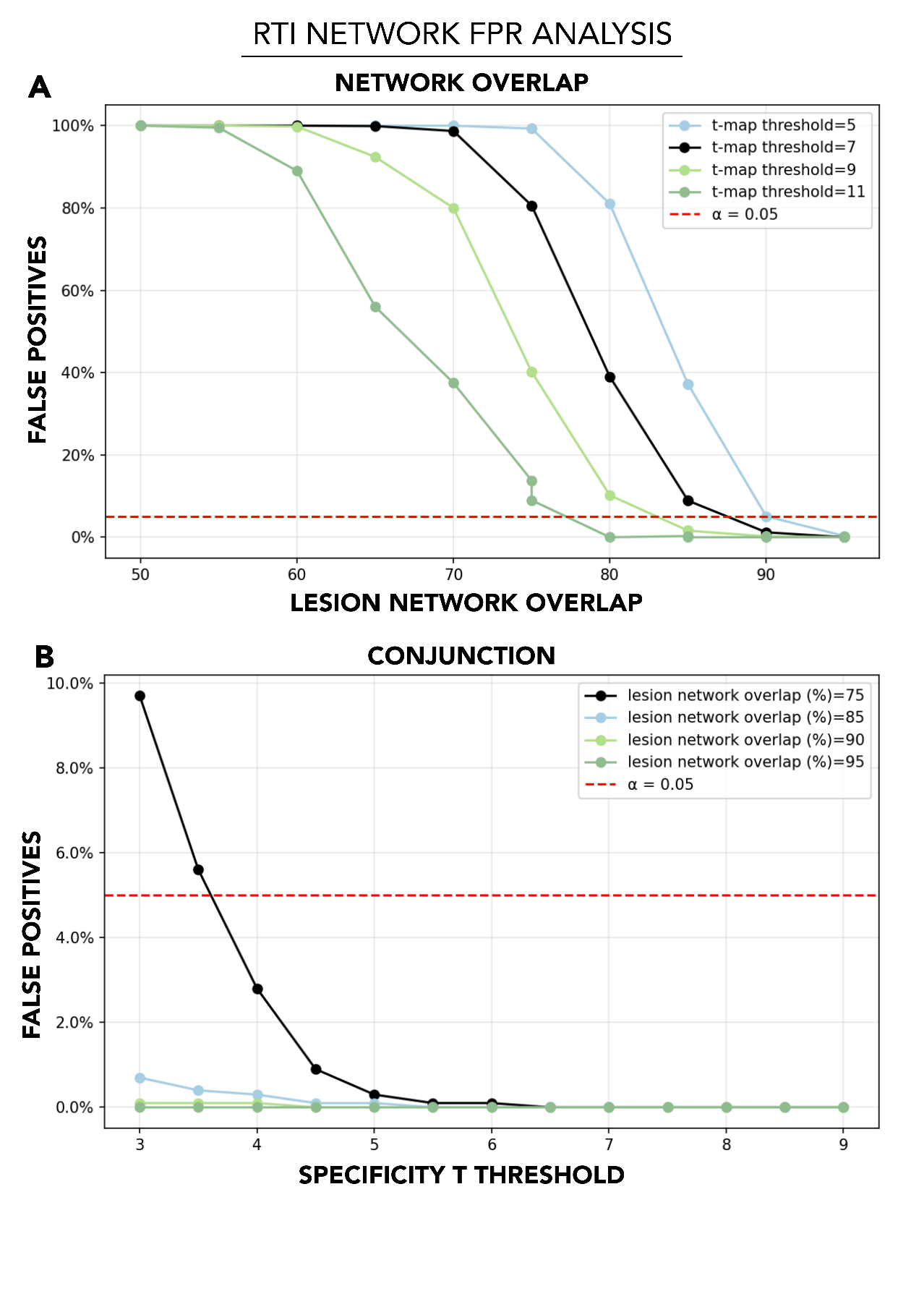

**Supplementary Figure 6**. The RTI network encompasses the majority (92%) of lesion locations and includes the cerebellum, brainstem, posterolateral thalamus as well as occipito-temporal and parietal cortex.

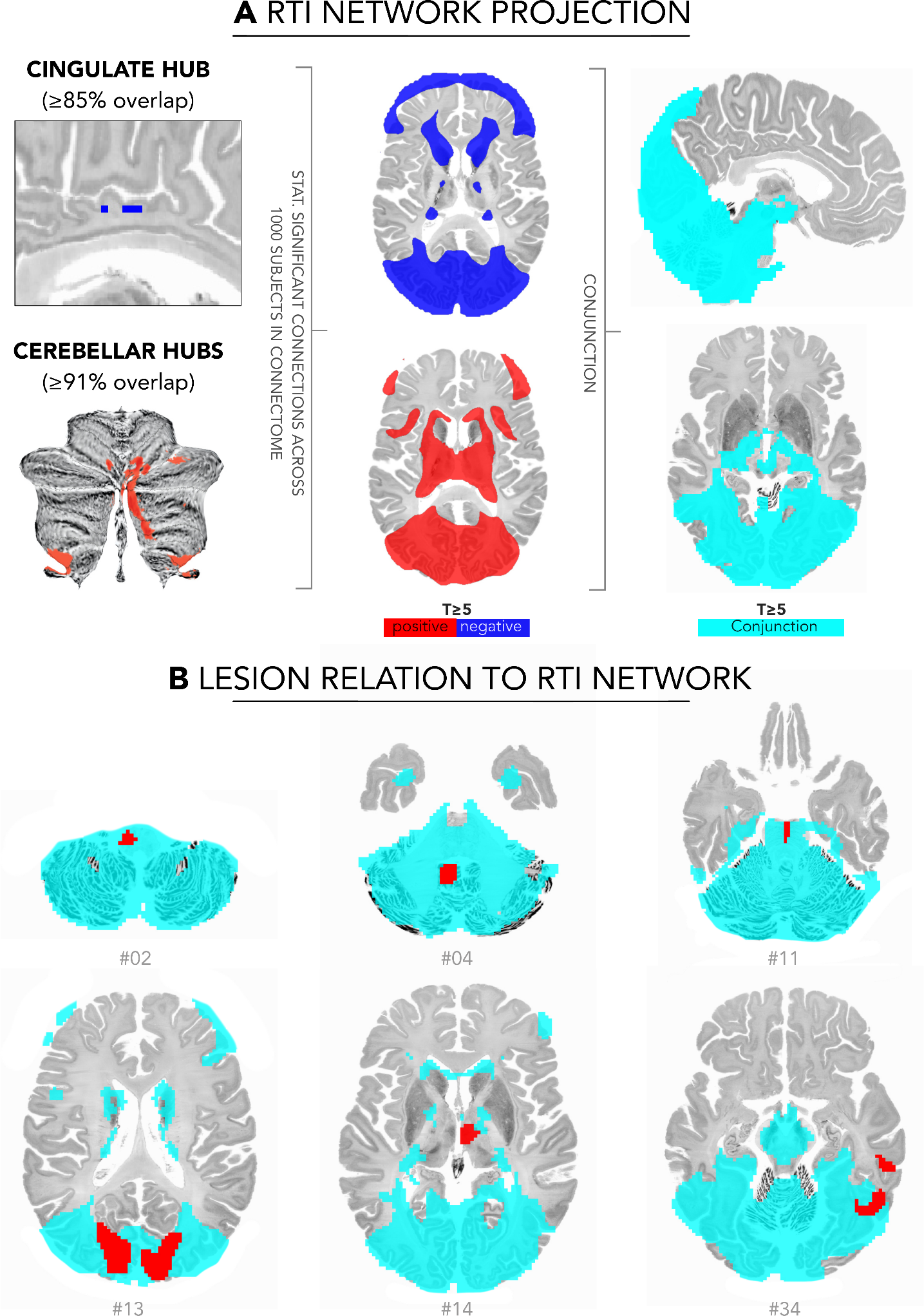

**Supplementary Figure 7**. A-B, Lesions causing milder disturbances of verticality perception show little overlap (max. 11%). However, a peak localized in the right posterior insula survives VLSM analysis, consistent with prior reports^4^. C, Disconnectome-based comparisons of RTI and milder verticality disturbances reveal only sparse differences after correction for multiple comparisons, while the unthresholded maps suggest two motifs: greater involvement of occipito-temporo–insular pathways in RTI, and greater disconnection of pontine and midbrain tegmental pathways in milder verticality disturbances.

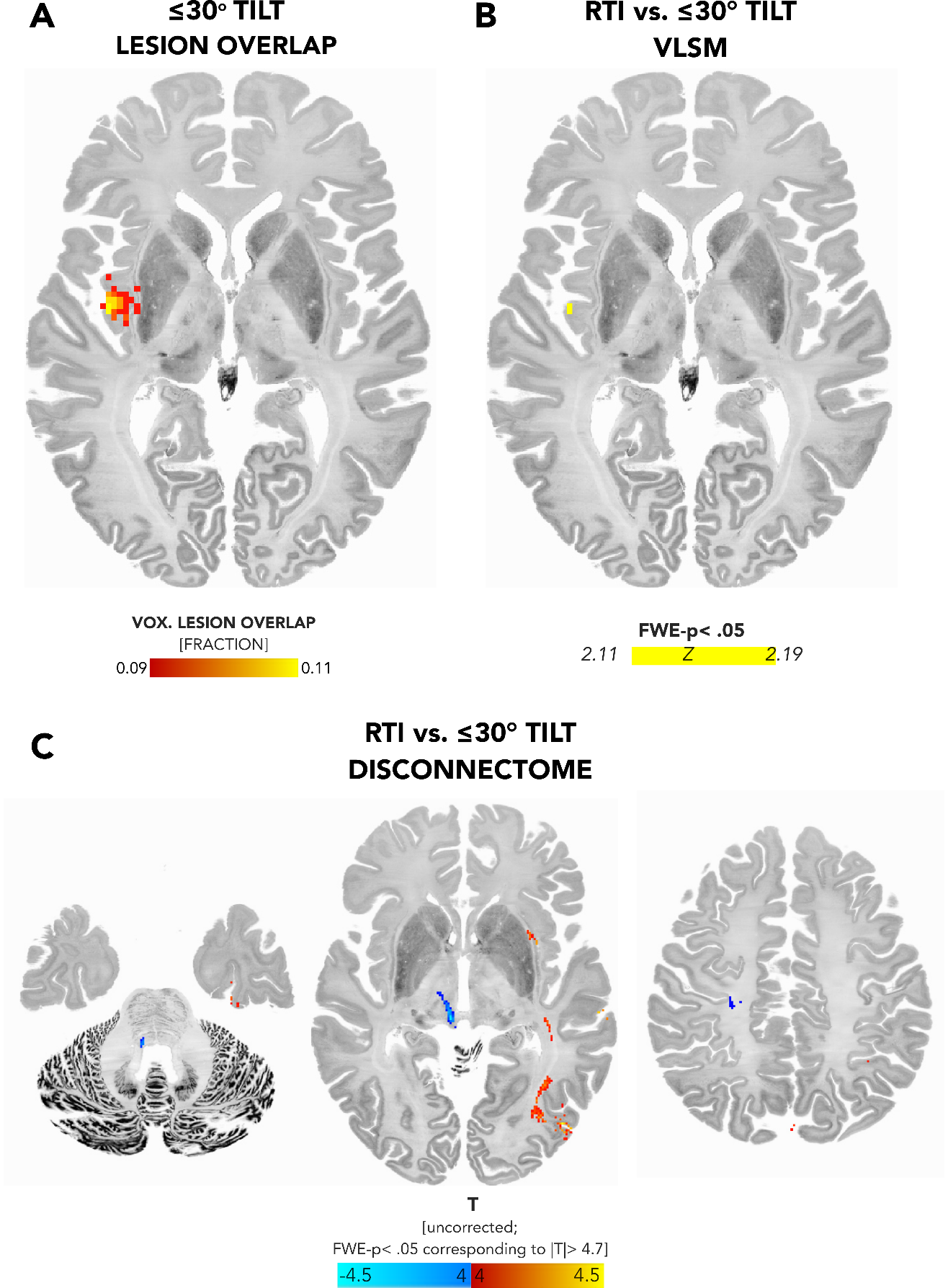

**Supplementary Figure 8**. A. Network mapping of lesions causing milder disturbances of verticality perception reveals hub regions mainly in the intermediate cerebellar lobules VI–VIII, with a topography notably distinct from that observed in RTI, and minimal overlap in the paravermal portion of lobule VI. B. Secondary clusters of significant differences between RTI and milder disturbances of verticality were identified in the bilateral posterior insulae (more negatively connected in RTI; region as defined by the Julich atlas^5^) and lateral occipital cortex (more positively connected in RTI, region defined by the AAL 3.0 atlas^6^).

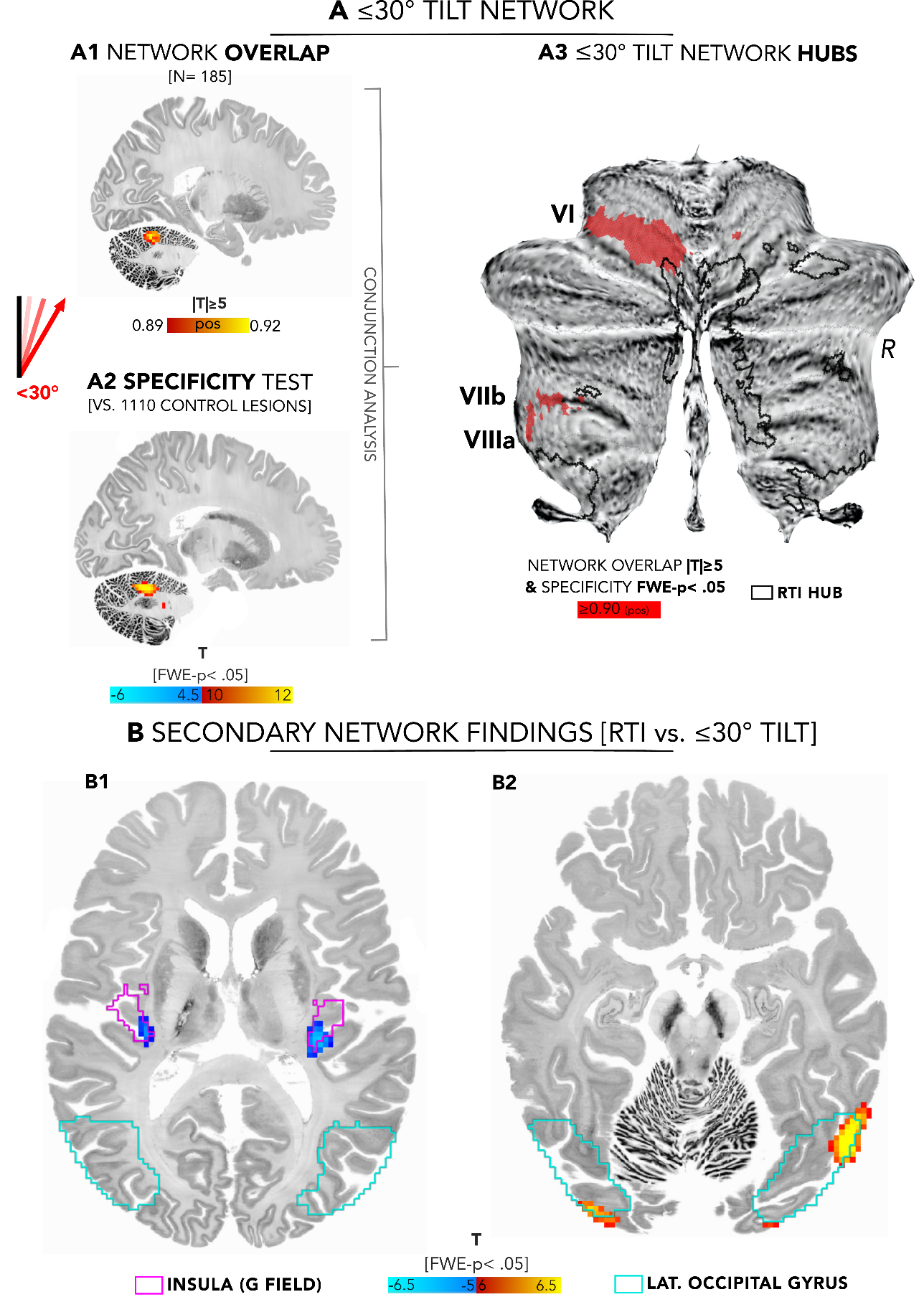

**Supplementary Figure 9.** Whole-brain connectivity analysis of direct electrical stimulation (DES) sites eliciting vestibular sensations. Left: Network overlap map showing regions significantly and consistently connected to vestibular DES sites (n= 57), thresholded at |T| ≥ 5 (FWE-p <. 05). Right: Voxel-wise specificity analysis comparing connectivity maps from vestibular DES sites with all remaining non-vestibular DES sites (n = 452), showing preferential connectivity of vestibular DES sites to the mid-cingulate cortex, extending into the adjacent precuneus (voxel-wise FWE-corrected p< .05). Black outlines indicate lesion-derived RTI hub regions.

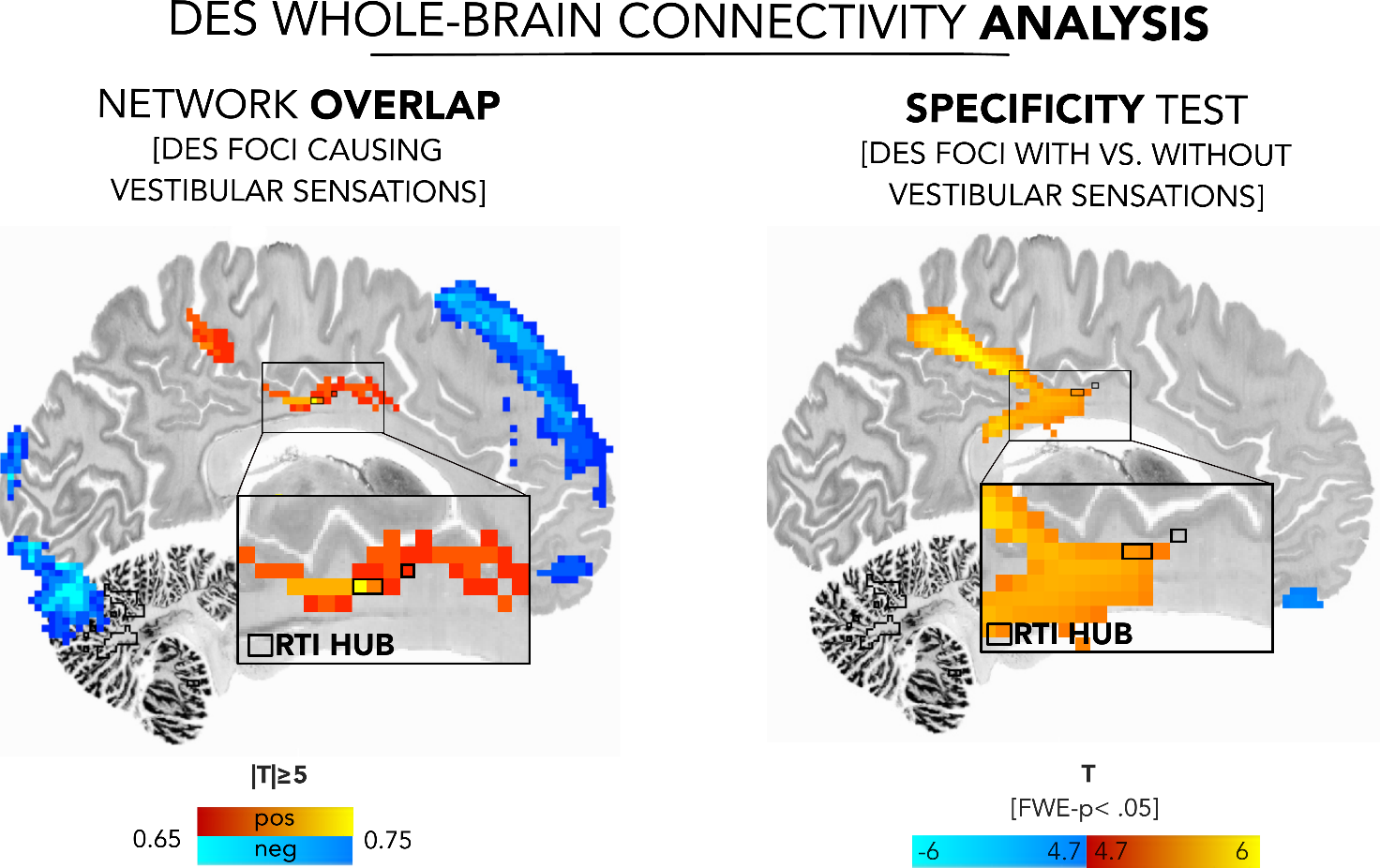
